# ECG classification with convolutional neural networks demonstrates resilience to sex-imbalances in data

**DOI:** 10.1101/2025.08.31.25334560

**Authors:** Diantha JM Schipaanboord, Floor van der Zalm, René van Es, Melle Vessies, Rutger R van de Leur, Klaske R Siegersma, Pim van der Harst, Hester M den Ruijter, N Charlotte Onland-Moret, Wouter AC van Amsterdam, the IMPRESS consortium

**Affiliations:** Laboratory of Experimental Cardiology, University Medical Center Utrecht, Utrecht University, Utrecht, the Netherlands; Julius Center for Health Sciences and Primary Care, University Medical Center Utrecht, Utrecht University, Utrecht, the Netherlands; Central Diagnostic Laboratory, University Medical Center Utrecht, Utrecht University, Utrecht, the Netherlands; Department of Cardiology, Division Heart and Lungs, University Medical Centre Utrecht, Utrecht University, Utrecht, the Netherlands

## Abstract

**Background:** Many ECG-AI models have been developed to predict a wide range of cardiovascular outcomes. The underrepresentation of women in cardiovascular disease studies has raised concerns if these models are equally predictive in women as compared to men. We tested the effect of sex-imbalance in training datasets on predictive performance of ECG-AI models, investigating imbalance in representation (ratio women-to-men), as well as in outcome prevalence, and percentage of misclassification.

**Methods:** We used a dataset containing raw 12-lead ECGs (n = 474,006) of 181,755 individuals who visited the University Medical Center Utrecht at any of the non-cardiology departments between July 1997 and August 2023 and sampled a sex-balanced dataset (n = 165,156) including only one ECG per individual. Multiple deep convolutional neural networks were trained to predict four outcomes; left bundle branch block, Long QT Syndrome, left ventricular hypertrophy or ECGs classified as ‘abnormal’ by a physician. Using subsampling, we simulated scenarios of sex-imbalance in representation (n_scenario_=5) for all outcomes and disease prevalence (n_scenario_=5), both representation and disease prevalence (n_scenario_=20) and disease misclassification (n_scenario_=7) for ‘abnormal’. Model performance was evaluated per scenario using area under the receiver operating characteristic curve (AUC) and smooth expected calibration error (smECE) for women and men separately.

**Results:** Across all scenario’s, the AUC remained stable, with small absolute differences between women and men for sex-imbalance in representation (ΔAUC: [0.002-0.025]), in disease prevalence (ΔAUC: [0.01-0.02]), in scenarios of both representation and disease prevalence (ΔAUC: [0.003-0.039]), and in outcome misclassification (ΔAUC: [0.007-0.077]). Only when disease prevalence in train and test data was sex-imbalanced, we observed differences in calibration error between sexes (max ΔsmECE: 0.26), with similar patterns for women and men.

**Conclusion:** The neural networks in this study demonstrated resilience to sex-imbalance in training ECG data.

**Graphical abstract:** 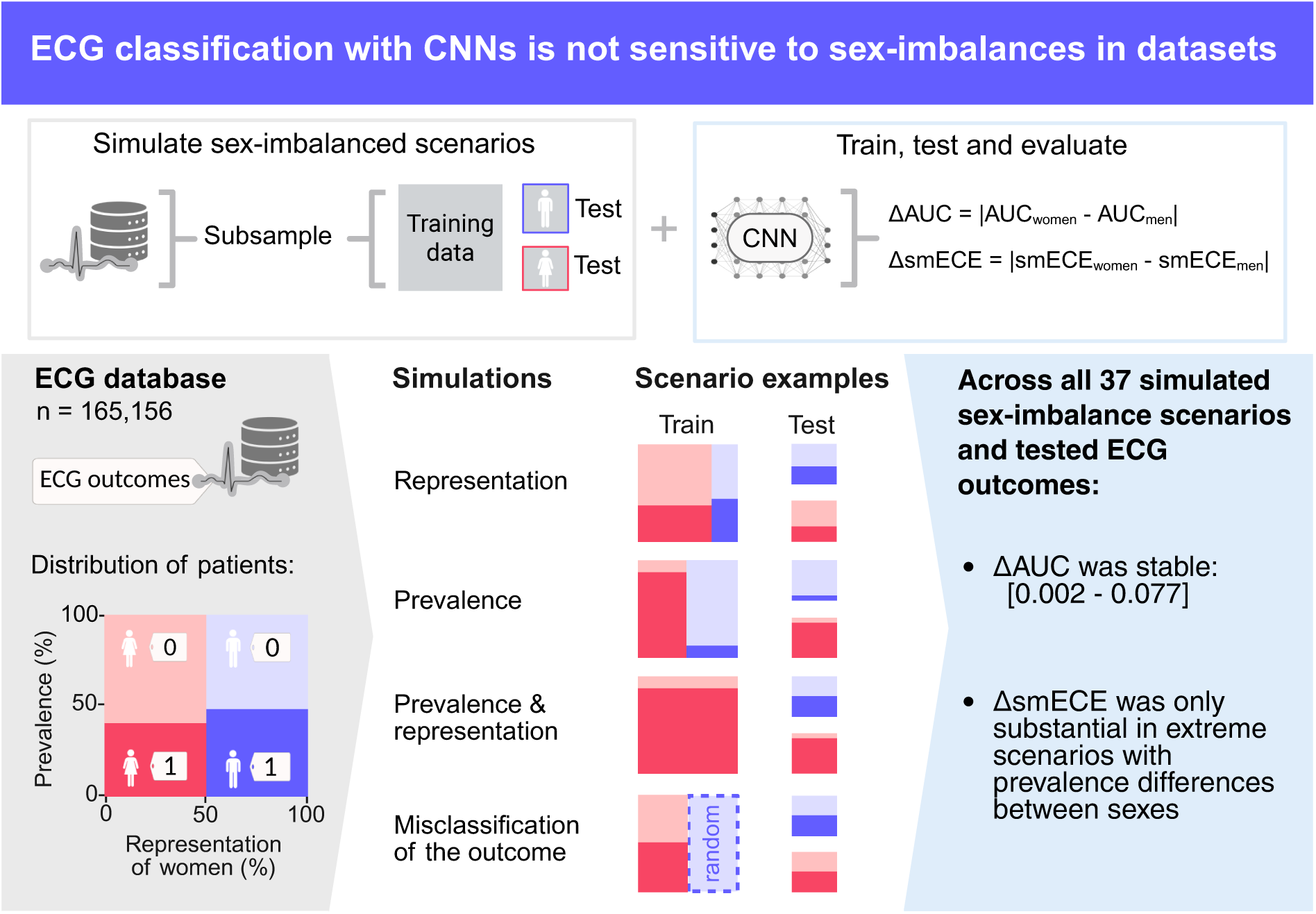

Graphical summary of the study methodology and results showing that ECG classification with convolutional neural networks is not sensitive to sex-imbalances in datasets. AUC = Area under the receiver operating curve; smECE = smooth expected calibration error. Created in BioRender. Meijer, I. (2025) https://BioRender.com/nxkwvoi.

## Introduction

The application of AI-based methods is increasing, and in the last decade numerous AI ECG-based prediction models for a broad range of cardiovascular outcomes have been developed.^1,2^ These models include deep learning models developed on a wide variety of datasets.

Women with cardiovascular disease are known to be underrecognized, underdiagnosed and undertreated.^3^ In datasets used for ECG deep learning, women are occasionally underrepresented^4–9^, while sex representation and sex-stratified performance is frequently unreported, allowing potential sex biases to go unnoticed.^10–16^ The implementation of biased algorithms may perpetuate healthcare disparities and exacerbate existing inequalities.^17^

We investigated whether the sex-balance in the training data used for the development of ECG prediction models using Convolutional Neural Networks (CNN) affects the predictive performance of these models. For this purpose, we created train and test datasets with sex-imbalance in representation (i.e. the percentage of women in the training set), prevalence of the outcome in women and men, and the combination of both by subsampling a large ECG dataset. Using these datasets, we trained CNN prediction models and assessed their predictive performance in women and men separately. In addition, we studied the effect of sex-dependent outcome misclassification on the predictive performance in women and in men.

## Methods

### Study design and population

For this study we used raw 12-lead ECGs (n=472,006) from 181,755 individuals who visited the University Medical Center Utrecht (UMC Utrecht) at any of the non-cardiology departments between July 1997 and August 2023. Most ECGs were recorded using the General Electric MAC 5500 (48.5%), MAC 5000 (32.9%), or MAC V (11.9%) resting ECG acquisition and analysis systems (GE Healthcare, Chicago, IL, USA), with a sampling frequency of 500 Hz (98.4%) or 250 Hz (1.6%).

The dataset included demographic features, such as sex and year of birth, extracted from hospital records and (diagnostic) ECG labels. Each ECG can have multiple labels, derived from free-text notes by physicians educated in ECG interpretation and annotation during cardiology residency. Physicians had access to patient demographics, medical records, previous ECGs, and computer-calculated conduction intervals. In addition to disease-specific annotations, the notes could include a general assessment, like annotating an ECG as ‘abnormal’. These annotations were extracted and structured using free-text mining software, resulting in binary diagnostic outcomes.^18^ All data were pseudo-anonymized.

We opted to predict diseases with known sex differences in ECG patterns and the established or suggested use of sex-corrected diagnostics criteria, resulting in the inclusion of Left Bundle Branch Block (LBBB), Long QT Syndrome (LQT) and left ventricular hypertrophy (LVH) as outcomes.^19–21^ We also selected the label ‘abnormal’ as an outcome because of its relatively high prevalence in the study population, allowing us to create scenarios with extreme imbalance in the predicted outcome without drastically reducing the number of ECGs. The most common additional labels of ECGs labelled as ‘abnormal’ were: sinus rhythm (62%), ST-T abnormalities (39%), a left heart axis (14%), LVH (11%), or atrial fibrillation (10%).

We excluded individuals with missing data on sex, year of birth, diagnostic outcomes used in our study, ECG sampling frequency, number of ECG leads, ECG acquisition date or device (n=14) and those of whom the ECG had a number of leads other than 8 (n=7) to maintain uniformity. Additionally, we excluded individuals with an age <18 years (n=4,994) or >120 years (n=2) at ECG acquisition. For individuals with multiple ECGs, one ECG was randomly selected. Finally, we randomly selected an equal number of men and women, resulting in a dataset of 165,156 ECGs (50% women) for further analyses (Figure 1).

**Figure 1:**
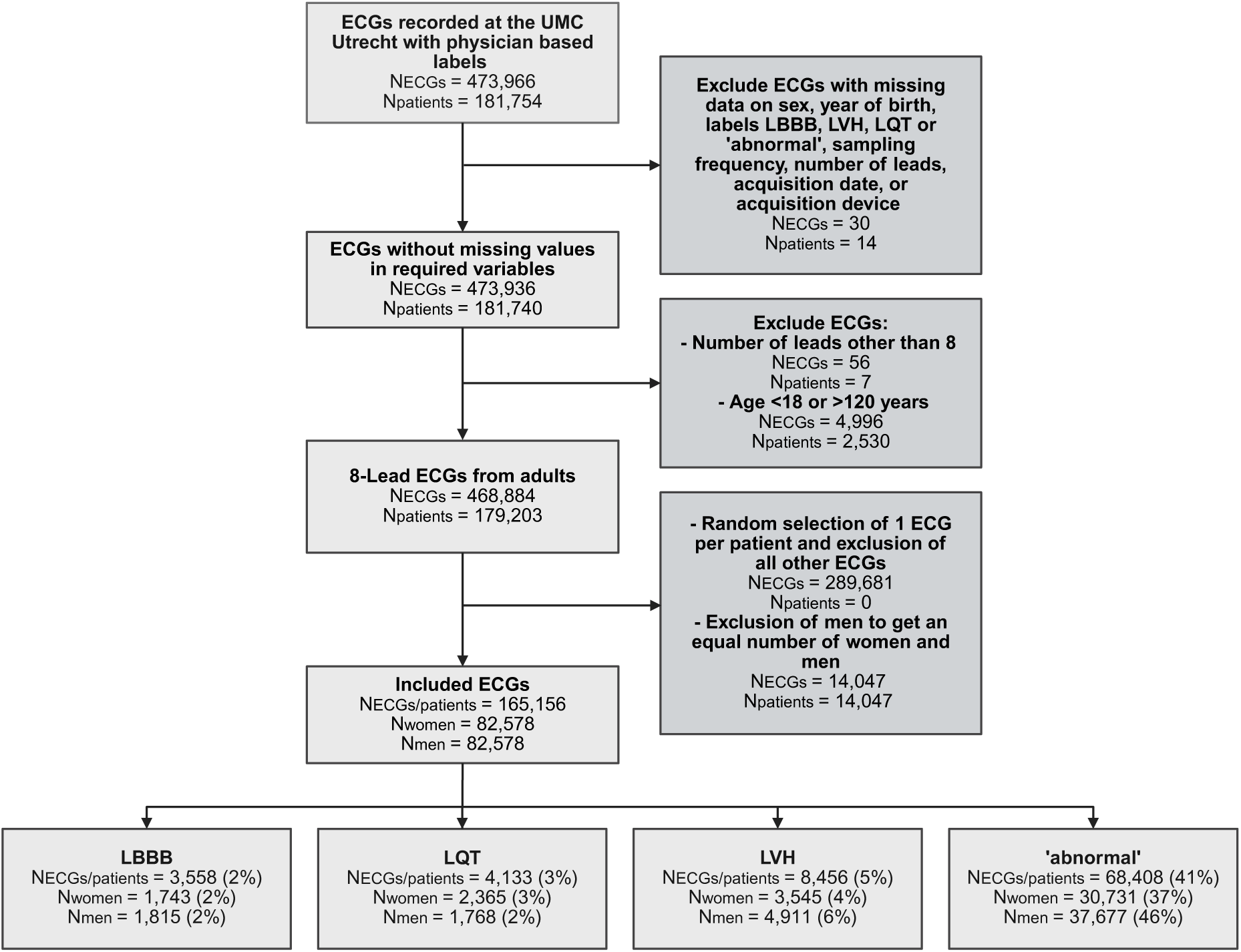
Flowchart of patient selection. LBBB = Left bundle branch block; LVH = Left ventricular hypertrophy; LQT = Long QT Syndrome; UMC Utrecht = University Medical Center Utrecht. Created in BioRender. Meijer, I. (2025) https://BioRender.com/lf8d4ue.

### Model input

We used the median ECG beats of lead 1, lead 2, and V1 to V6 as generated by the MUSE ECG system (MUSE version 8, GE Healthcare, Chicago, IL, USA) as input to train, validate and test the model.^18^ ECGs recorded at 250 Hz were resampled to 500 Hz using linear interpolation.

### CNN architecture and hyperparameters

The architecture of the CNN was previously proposed, described and optimized in prior studies.^13,22–25^ In contrast to previous studies, training was stopped when the Binary Cross Entropy (BCE) loss on the validation dataset did not improve by more than 0.001 for 10 consecutive epochs, with a maximum of 50 epochs. A threshold value of 0.5 was applied to the probabilities to determine class labels for the performance metrics. The ECGxAI package (version 0.1.1) was used to import ECGs and set up the training, testing and validation.^13^ This package utilized the deep learning framework PyTorch (version 2.1.0) and PyTorch-Lightning (version 1.5.10) to fit and evaluate the model.^26^ All analyses were conducted using Python (version 3.8.10).^27^

### Simulating sex-imbalances

We simulated four types of sex-imbalance in training data: (1) sex-imbalance in representation, (2) sex-imbalance in outcome prevalence, (3) sex-imbalance in both representation and outcome prevalence, and (4) sex-imbalance in misclassification (i.e. mislabelled outcomes in the training sets). Each simulation contained scenarios with varying degrees of sex-imbalance. We used an 1:1 women-to-men ratio in all test sets to ensure balanced evaluation across sexes.

The subsampling steps for each scenario are detailed in Methods S1. Briefly, we first determined the maximum number of ECGs available for each simulation and outcome (e.g. simulation 1; LBBB) based on our original dataset. For each scenario, we calculated sample sizes for women and men with and without the outcome, considering a 50:20:30 train:validation:test ratio and necessary sex representation and outcome prevalence. We then subsampled the ECGs, split the data into training, validation, and test sets, and repeated the process five times (5-fold stratified cross-validation). Models were trained using an identical CNN architecture, with performance evaluated by sex. Sample sizes varied by simulation and outcome but were similar across scenarios within a simulation.

### Simulation 1: Sex-imbalance in representation

First, we evaluated the effect of sex representation in the training data on the sex-stratified model performance in five scenarios (Figure 2A). We subsampled with the following proportions of women in the training and validation datasets: 1.00, 0.75, 0.50, 0.25, and 0.00. In all datasets the sex-specific outcome prevalence was equal to the original dataset. This simulation was done for outcomes LBBB, LQT, LVH, and ‘abnormal’.

**Figure 2:**
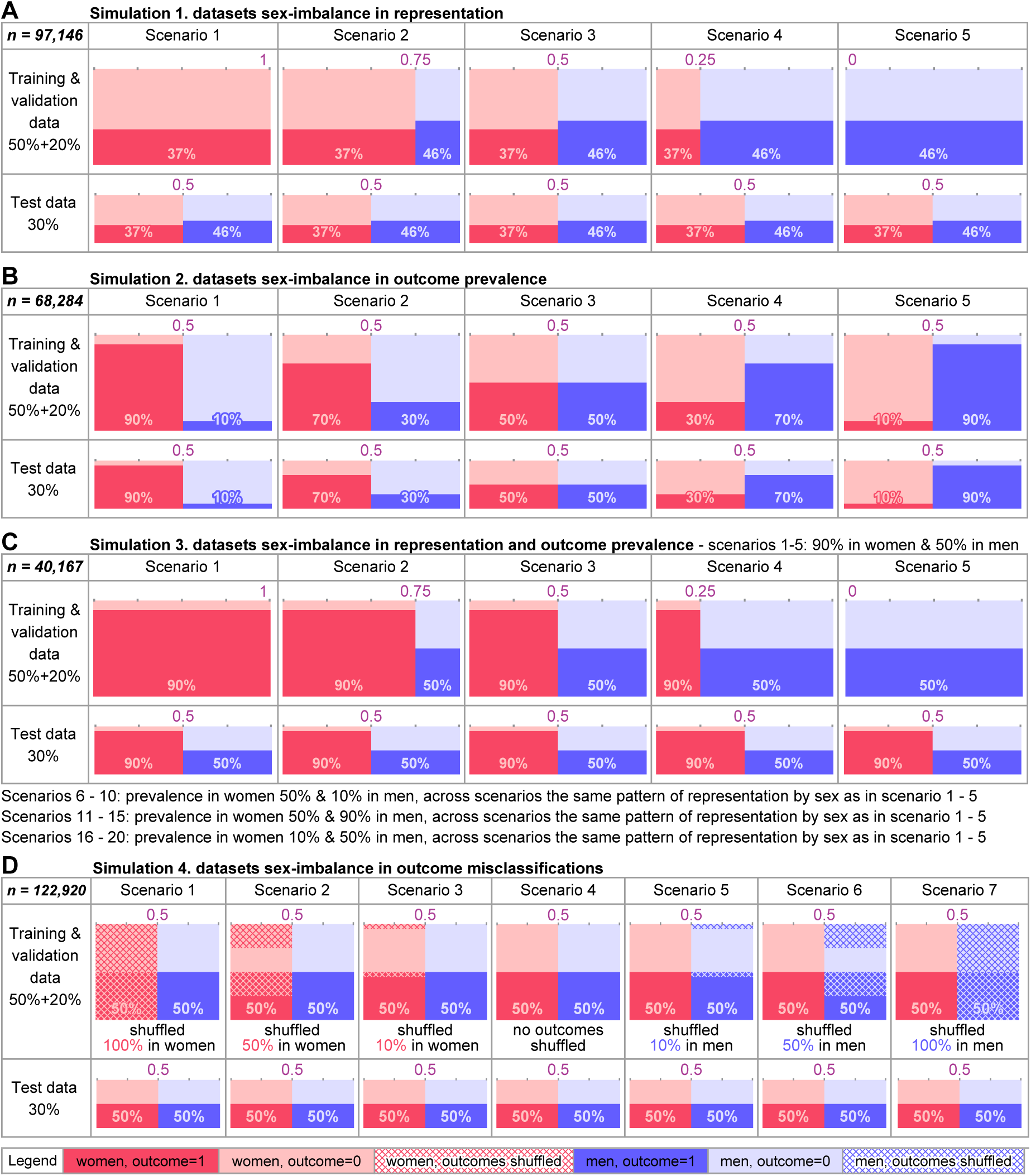
Simulation datasets. Visualization of training, validation and test data for the scenarios predicting outcome ‘abnormal’ of: **A)** simulation 1: sex-imbalance in representation, **B)** simulation 2: sex-imbalance in outcome prevalence, **C)** simulation 3: sex-imbalance in representation and outcome prevalence, and **D)** simulation 4: sex-imbalance in outcome misclassification. Each training & validation dataset shows the proportion of women in purple and the prevalence of the predicted outcome by sex. The total number of individuals used in each simulation is depicted per simulation in the top left corner.

### Simulation 2: Sex-imbalance in outcome prevalence

In the second simulation, we assessed the effect of sex-differences in outcome prevalence on the sex-stratified model performance, again in five scenarios (Figure 2B). We only used the label ‘abnormal’ as outcome because of its high prevalence in the original dataset. We subsampled datasets with the following women-to-men outcome prevalences in the training, validation and test data: 90%:10%, 70%:30%, 50%:50%, 30%:70%, and 10%:90%. In each scenario, the proportion of women and men was subsampled to be equal.

### Simulation 3: Sex-imbalance in both representation and outcome prevalence

Next, we constructed 20 scenarios combining sex-imbalances in representation and outcome prevalence to assess their joint effect on sex-stratified model performance (Figure 2C). Again, we only used ‘abnormal’ as outcome. We first created four datasets with the following women-to-men outcome prevalences: 90%:50%, 10%:50%, 50%:90% and 50%:10%, by selectively subsampling from the original dataset. For each prevalence-imbalanced dataset, we created training, validation, and test datasets by subsampling the proportion of women in training and validation to 1.00, 0.75, 0.50, 0.25, or 0.00, as in the first simulation.

### Simulation 4: Sex-imbalance in outcome misclassifications

At last, we examined the effect of varying degrees of sex-dependent misclassification of the predicted outcome ‘abnormal’ in the training data on the sex-stratified model performance (Figure 2D). Initially, we created a balanced subsample by sex and outcome. Next, we simulated one reference and six misclassification scenarios by randomly shuffling a percentage of outcomes (women:men): 100%:0%, 50%:0%, 10%: 0%, 0%:0% (reference), 0%:10%, 0%:50%, 0%:100%.

### Statistical analysis

We assessed the performance of each model sex-stratified based on its discriminatory performance using the area under the receiver operating characteristic curve (AUC) and its calibration performance using the smooth expected calibration error (smECE), a hyperparameter-free calibration measure^28^. For all scenarios in a simulation, sex-stratified AUC and smECE were depicted in scatterplots, with error bars representing the 95% confidence interval obtained through bootstrapping (n=1,000) of the sex-stratified performance metrics. When notable calibration differences were observed between women and men, we generated calibration plots to assess over- and underprediction^28^.

### Sensitivity analysis

ECG prediction models are often developed on smaller datasets than ours.^29,30^ We, therefore, performed a sensitivity analysis by repeating the third simulation on two smaller datasets (train + validation: n=12,800 ECGs; n=1,280 ECGs). Second, we previously demonstrated a high AUC for predicting sex based on ECGs (0.96, 95% CI: 0.96-0.97) ^24^ and therefore repeated the third simulation using only patients where sex was correctly predicted by a CNN sex classification model, to assess if this could influence the difference in predictive performance in women and men (Method S2).

## Results

In total, 165,156 patients (50% women) were included, with a median age of 62 years (IQR: 49-72). Women and men did not differ in age or in year of ECG acquisition (Table S1). The conduction times of the ECGs were also similar between sexes, except for a shorter median QRS duration in women (88 ms (IQR: 80-96) vs. 98 ms (IQR: 88-106) in men) (Table S1). In women and men, the prevalence of LBBB was 2% vs 2%, LQT 3% vs 2%, and LVH 4% vs 6%, respectively (Figure 1). The ECG label ‘abnormal’ was highly prevalent in our study population (41%) and differed between women and men (37% vs 46%, respectively) (Figure 1).

### Simulation 1: Sex-imbalance in representation

Each dataset in simulation 1 (Figure 2A) consisted of around 48,574 ECGs in the training set, 19,429 ECGs in the validation set, and 29,142 ECGs in the test set (Table S2-S5). In general, the AUCs were high (range 0.878 to 0.997) and remained stable across all scenarios for all outcomes in both sexes (Figure 3A). The calibration error (smECE) was also stable across scenarios for each of the four outcomes, with minimal smECEs for LBBB (range 0.004 to 0.011), LQT (range 0.016 to 0.023), and LVH (range 0.009 to 0.024) and slightly higher smECEs for ‘abnormal’ (range 0.037 to 0.075) (Figure 3B). There were no differences in the smECEs between sexes, with the exception in the scenario where only women were used to train the model to predict the label ‘abnormal’ where the smECE was slightly higher in men than in women (0.075 vs 0.05). The test metrics and confidence intervals for all scenarios are also given in Tables S2B, S3B, S4B and S5B.

**Figure 3:**
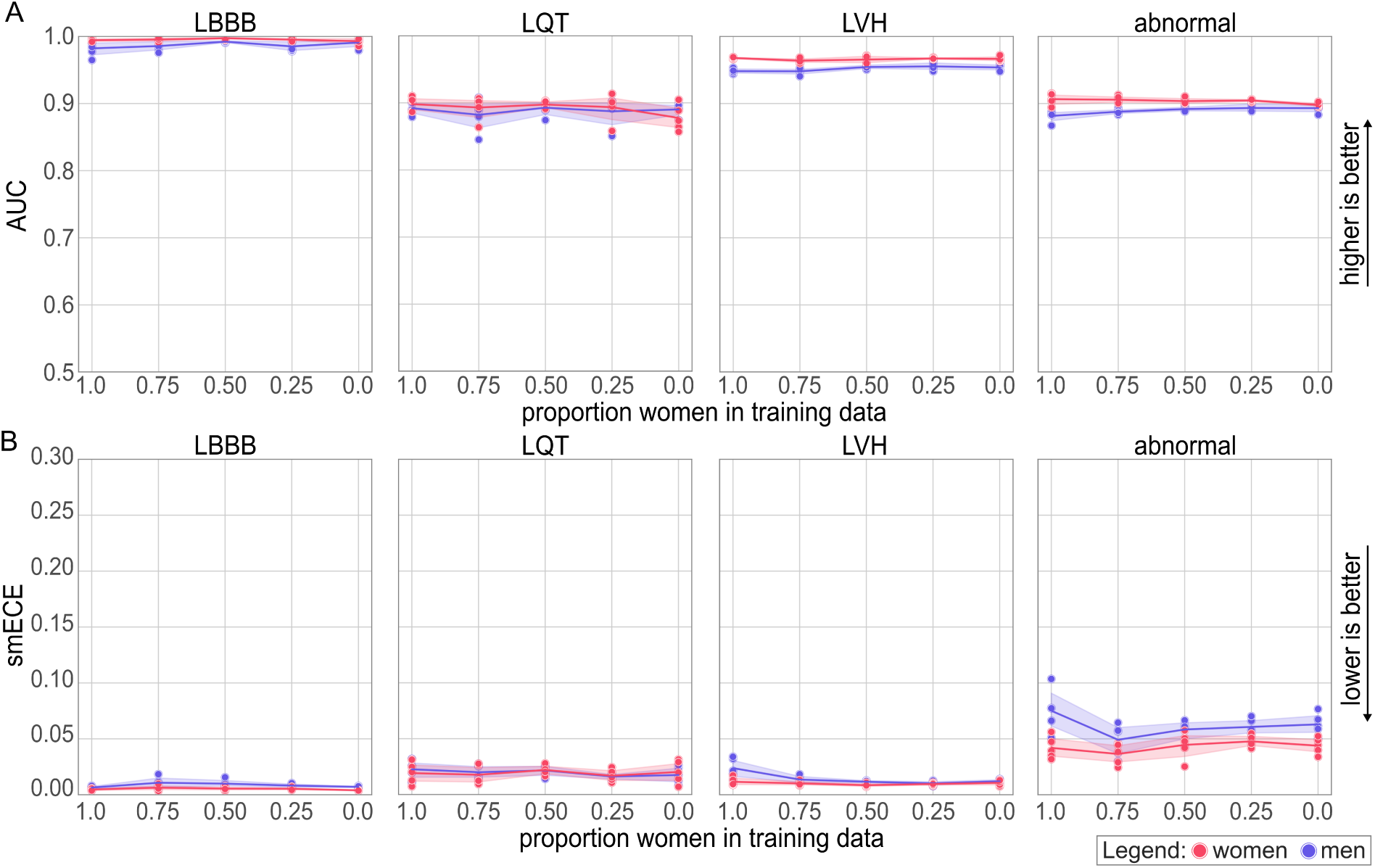
Sex-stratified model performance in simulated scenarios of sex-imbalance in representation. **A)** AUC of models that predict left bundle branch block (LBBB), Long QT Syndrome (LQT), left ventricular hypertrophy (LVH) and ‘abnormal‘. **B)** Calibration error of models that predict LBBB, LQT, LVH and ‘abnormal‘. Each dot depicts metrics of a single fold in women or in men, the line represents their average and the coloured area shows the bootstrapped 95% confidence interval.

### Simulation 2: Sex-imbalance in outcome prevalence

In simulation 2, around 34,144 ECGs were included in the training sets, around 13,656 ECGs in the validation sets, and around 20,486 ECGs in the test sets (Figure 2B and Table S6). We only used label ‘abnormal’ as the outcome.

The AUC was highest in both women and men when the prevalence of the outcome was equal for both sexes (50:50) (Figure 4A; women: 0.901, men: 0.889). Interestingly, larger prevalence differences between sexes resulted in a lower AUC in both sexes. But overall, the AUCs remained high (minimum AUC women vs men: 0.856 vs 0.835) (Figure 4A). The calibration error (smECE), however, was affected in a sex-specific way across the scenarios. When the prevalence was similar in women and men, the smECE was also most similar (smECE women vs men: 0.063 vs 0.061) (Figure 4B, Table S6). When the prevalence of the outcome started to differ between the sexes, the calibration error was systematically larger in the sex with the highest prevalence (smECE women vs men: ratio 90:10 = 0.177 vs 0.072; ratio 70:30 = 0.150 vs 0.045; ratio 30:70 = 0.028 vs 0.074; ratio 10:90 = 0.074 vs 0.106). The calibration error was slightly more affected by differences in prevalence in women than in men (Figure 4B and Table S6).

**Figure 4:**
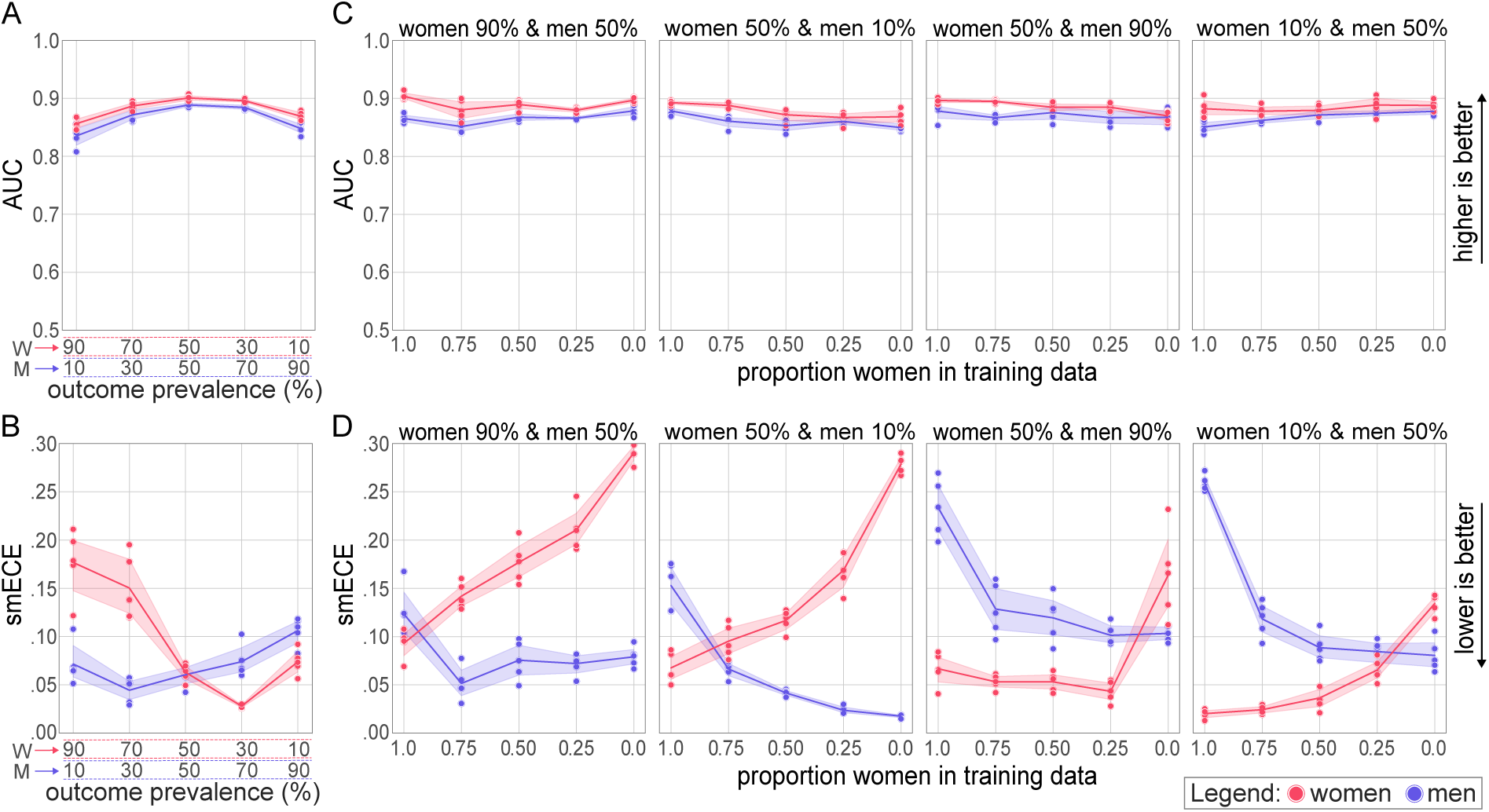
Sex-stratified model performance in simulated scenarios of sex-imbalance in outcome prevalence and sex-imbalance in both representation and outcome prevalence. **A)** AUC and **B)** calibration error of the models that predict ‘abnormal’ in scenarios sex-imbalance in outcome prevalence (simulation 2), **C)** AUC and **D)** calibration error of the models that predict ‘abnormal’ in scenarios of sex-imbalance in outcome prevalence and representation (simulation 3), with scenarios with equal outcome prevalences grouped in subplots. Each dot depicts metrics of a single fold in men or in women, the line represents their average and the coloured area shows the bootstrapped 95% confidence interval. W = Women, and M = Men.

### Simulation 3: Sex-imbalance in both representation and outcome prevalence

In the third simulation (Figure 2C), we selected around 20,084 ECGs in each training set, 8,034 ECGs in each validation set, and 12,049 ECGs in each test set (Table S7-S10). Again, only the outcome ‘abnormal’ was used. Again, the AUC was high and remained stable (range 0.850 to 0.904) across all scenarios tested (Figure 4C) but was consistently higher in women compared to men (max difference 0.039). Similar to simulation 2, the calibration error (smECE) was consistently higher in the sex with a higher prevalence of the outcome, except in the scenario in which only individuals were included of the sex with the highest prevalence. In that situation the smECE was lower in individuals of that sex (Figure 4D). Calibration plots show that the model consistently under-predicted outcomes for the sex with higher outcome prevalence (Figure S1). But if the outcome prevalence was low in for instance women, the model overpredicted the outcome in women when women were underrepresented in the training set.

### Simulation 4: Sex-imbalance in outcome misclassifications

To simulate the effect of differential misclassification of the outcome between the sexes we included around 61,460 ECGs in the training set, 24,584 ECGs in the validation set and, 36,876 ECGs in the test set (Figure 2D, Table S11). Again, the predictive performance was high in both women and men with AUCs ranging between 0.80 and 0.91. When misclassification of the outcome was introduced in men, the predictive performance decreased in men and a similar effect was seen for women (Figure 5A). However, this decrease in AUC was small, with a maximum decrease in AUC of only 0.094. The calibration error was not affected by differential misclassification of the outcome but was slightly higher in women than in men (range 0.005 to 0.031) across all misclassification scenarios (Figure 5B).

**Figure 5:**
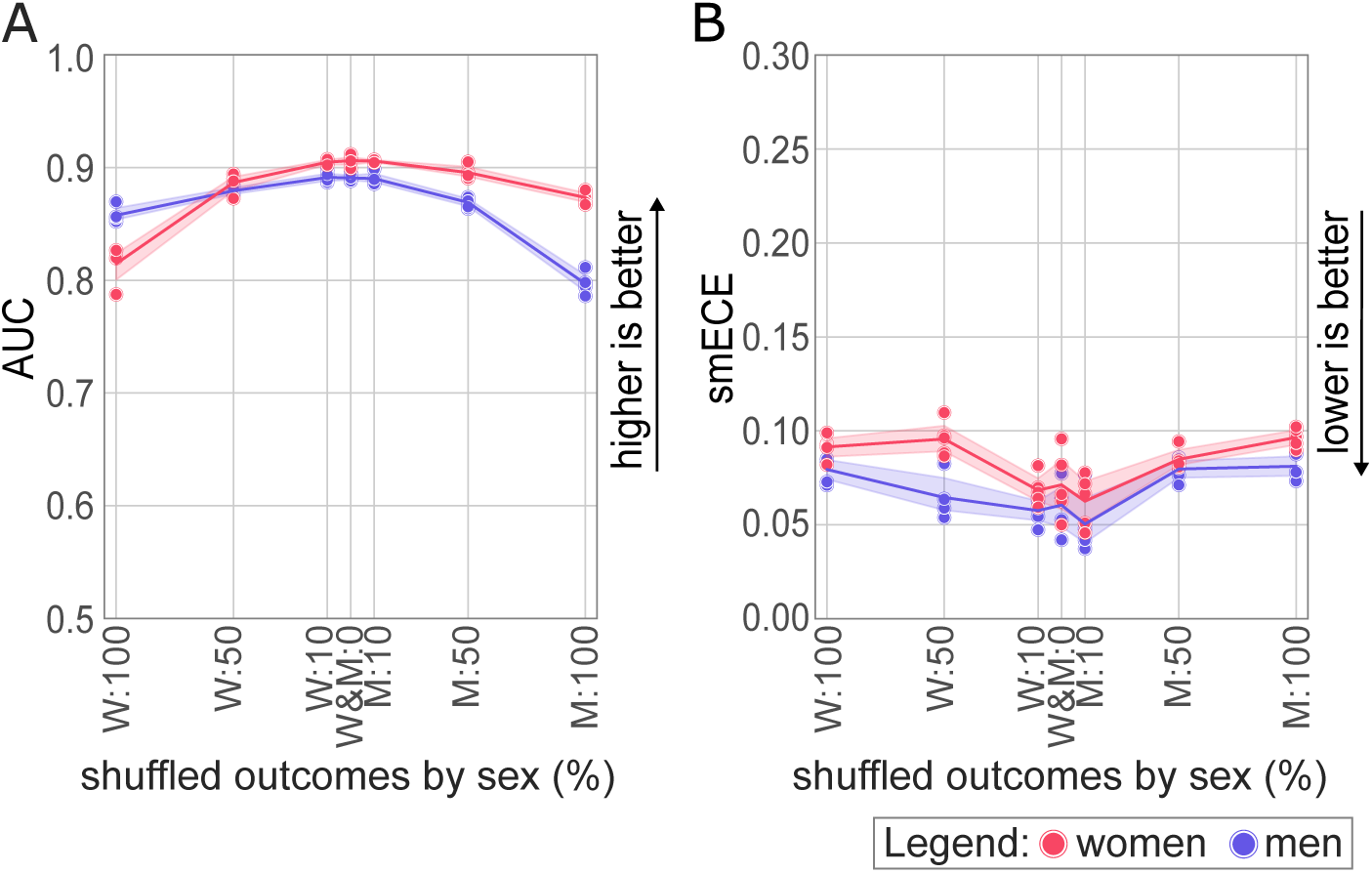
Sex-stratified model performance in simulated scenarios of sex-dependent outcome misclassification. Misclassifications were introduced by randomly shuffling certain percentages of the predicted outcomes in women or in men (simulation 4). Each dot depicts metrics of a single fold in men or in women, the line represents their average and the coloured area shows the bootstrapped 95% confidence interval. **A)** AUC and **B)** calibration error of the models that predict ‘abnormal’ in scenarios sex-imbalance in outcome prevalence. W = women, M = men.

### Sensitivity analysis

To assess the influence of sample size on predictive performance in women and men, we repeated the third simulation using two smaller datasets. Smaller sample size did not affect AUCs and smECEs when compared to the third simulation (Figure S3).

We also assessed the impact of sex-misclassified patients by repeating the third simulation with only those whose sex was correctly predicted by a CNN sex classification model. Again, the AUCs and smECEs were similar to those of the third simulation (Figure S4), suggesting that our findings were unaffected by sample size or the presence of ECG-based sex-misclassified patients.

## Discussion

We showed that CNN models that predict disease using ECGs as input are robust to variation in representation and outcome prevalence between sexes and to sex-dependent outcome misclassification. The model performance was stable across the various sex-imbalance scenarios and only diverged slightly in extreme and unrealistic scenarios, with similar results for both women and men for opposite-sex scenarios.

The stability in AUC indicates that the models are not overly dependent on sex-specific ECG-patterns when making disease predictions. The sensitivity analyses confirmed that our results were not influenced by the large sample size or by the fact that CNNs can classify sex based on ECGs with high accuracy.^24^

Our results align with other research demonstrating a CNN model trained to detect low left ventricular ejection fraction of ECGs exhibited consistent performance across racial/ethnic subgroups, even though a separate CNN model was able to discern these racial/ethnic subgroups.^31^

While AUC remained stable across scenarios, we observed that sex differences in disease prevalence influenced the smECEs (simulation 2-3), with higher smECEs for the sex with higher outcome prevalence. We showed that this could be explained by underprediction. In these simulations, the model predicted whether an ECG was labelled ‘abnormal’ by a physician, a label that includes various ECG abnormalities. Calibration was likely affected by the more homogeneous non-diseased group, leading to higher miscalibration (i.e. higher smECEs) in the sex with higher prevalence of ‘abnormal’ ECGs in the test data. Overprediction occurred when the lower-prevalence sex was underrepresented in training, suggesting that the model may rely more on overall disease prevalence than on sex-specific ECG features.

### Strengths and limitations

A strength of our study is the use of extensive simulations and additional sensitivity analyses, which allowed us to thoroughly challenge and evaluate model performance under a wide range of sex-imbalances.

Second, our study focused on three commonly occurring outcomes (i.e. LBBB, LQT and LVH) and an outcome which is a collection of a wide range of ECG abnormalities. Given the consistent findings, especially on the relatively well sex-balanced datasets, similar results might be expected for deep learning ECG prediction models for other diseases. However, confirmation in independent studies is warranted.

Moreover, we evaluated model performance by both discrimination and calibration. This dual approach provides a more comprehensive understanding of the model’s behaviour across women and men, revealing both its ability to distinguish outcomes and the reliability of its predicted probabilities.

This study is a small step in the broader effort to assess and achieve algorithmic fairness. We only focused on ECG classification and CNN models, meaning that our finding of robust performance despite sex-imbalances may not be generalizable to other types of input data, such as clinical variables or images, or other machine learning applications. Furthermore, our results cannot be generalized to imbalances based on other characteristics, such as ethnicity. Unfortunately, these characteristics were not available to us.

Another limitation lies in the method used to simulate misclassification of ECG labels, which did not account for the underlying causes of potential misclassification. Misclassification is unlikely to be entirely random and may differ systematically between women and men. Nevertheless, our goal was to challenge the model to its extreme limits. By focusing on extreme scenarios, we have gained a clear understanding of potential trends and directions.

For future research, we recommend investigating whether our finding of robust model performance to sex-imbalances also holds across other diseases, other types of input data and alternative model architectures beyond CNNs. Furthermore, we recommend performing other subgroup analyses beyond sex and ethnicity, for instance age and cardiovascular risk factors, to increase insight in the generalizability and fairness of machine learning models.^32^

## Conclusion

Our research demonstrated that a CNN model that uses ECGs to predict disease is robust to sex-imbalance in representation and outcome prevalence and sex-dependent outcome misclassification in the training data. Future studies should clarify whether our results are generalizable to other types of data, other machine learning applications, and other subgroups.

## Data Availability

The data cannot be shared publicly. The code relevant to this work is available on a private GitHub repository (SexImbalanceAI) and can be accessed upon reasonable request.

## Funding

This project is part of the Dutch Cardiovascular Alliance Consortium IMPRESS (2020B004).

## Acknowledgements

We would like to acknowledge Qiao Zhou for having previously explored this question. We used AI-assisted technologies (ChatGPT, OpenAI) for code debugging and refinement and to enhance the readability and conciseness of the manuscript through linguistic suggestions.

## Conflicts of interest

The Department of Cardiology at UMC Utrecht may receive royalties in the future from sales of deep learning ECG algorithms developed by Cordys Analytics, a spin-off company. Additionally, RvE is the Chief Scientific Officer (CSO) and RvdL is Chief Medical Officer (CMO) and both are shareholders of Cordys Analytics. These affiliations and potential financial interests have been disclosed and are being managed in accordance with institutional policies.

## Notes

### Author Declarations

The University Medical Center Utrecht ethical committee approved this research.

