## Supplementary Material for "ECG classification with convolutional neural networks demonstrates resilience to sex-imbalances in data"

Method S1

Method S2

Table S1

Table S2

Table S3

Table S4

Table S5

Table S6

Table S7

Table S8

Table S9

Table S10

Table S11

Figure S1

Figure S2

Figure S3

Figure S4

### Method S1

The process of subsampling to simulate the sex-imbalanced datasets required the following steps:

**Step 1: Determine the maximum number of ECGs possible for a single simulation and a single outcome.**

To maintain consistency across all scenarios in a simulation, we calculated the maximum possible number of individuals (N) based on the dataset's properties and composition using the following formula:

$$N = \left\lfloor \frac{\min\left(\frac{N_{\text{women,outcome1}}}{\max(P_{\text{women}})}, \frac{N_{\text{women,outcome0}}}{(1-\max(P_{\text{women}}))}, \frac{N_{\text{man,outcome1}}}{\max(P_{\text{man}})}, \frac{N_{\text{man,outcome0}}}{(1-\max(P_{\text{man}}))}\right)}{FS_{max}} \right\rfloor \quad (1)$$

Where:

$N_{women,outcome1}$: $|\ \{p \in P \mid Sex(p) = Woman\ \wedge outcome(p) = 1\}\ |$

$N_{women,outcome0}$: $|\ \{p \in P \mid Sex(p) = Woman\ \wedge outcome(p) = 0\}\ |$

$N_{men,outcome1}$: $|\ \{p \in P \mid Sex(p) = Man\ \wedge outcome(p) = 1\}\ |$

$N_{men,outcome0}$: $|\ \{p \in P \mid Sex(p) = Man\ \wedge outcome(p) = 0\}\ |$

$FS_{max}$: maximum fraction of included measurements from one sex $= max(Fraction\_of\_sex\_in\_train) \times Fraction_{train\_and\_validation\_data} + 0.5 \times Fraction_{test\_data}$

$P_{woman}$: list of examined prevalence percentages of the selected ECG diagnostic outcome in $p \in P \mid Sex(p) = Female$

$P_{man}$: list of examined prevalence percentages of the selected ECG diagnostic outcome in $p \in P \mid Sex(p) = Male$

$N$: number of included ECG measurements for each fold for CNN training and testing.

For simulation 3, which required multiple prevalence scenarios, this formula was applied to all scenarios, and the smallest resulting number was used to ensure consistency across them.

**Step 2: Determine sample sizes for each dataset and group.**

We calculated the sample sizes for the training, validation, and test datasets for each scenario based on the necessary sex representation and outcome prevalence for that scenario, and the predefined ratio (50:20:30) between training, validation, and test datasets. This was done separately for the following four groups:

- Women with outcome = 1
- Women with outcome = 0
- Men with outcome = 1
- Men with outcome = 0

**Step 3: Subsample the ECG measurements for each group.**

The total number of ECG measurements was subsampled per group according to the calculated sample sizes for each scenario.

**Step 4: Split data into training, validation, and test sets.**

The subsampled data was split into training, validation, and test datasets. This process was repeated for each five folds.

### Method S2

For the second sensitivity analysis, we used the dataset of the third analysis (n=165,156). In total, 32% (n=52,849) was used to train and 8% (n=13,212) to validate a sex classification model, using an identical CNN architecture. We tested the sex classification model on the remaining 60% (n=99.095), resulting in a sex probability for each patient. A threshold value of 0.5 was applied to the probabilities to determine the sex class. The performance metrics of the sex classification model are shown in the table below. Thereafter, we repeated the third analysis on the full dataset previously used for testing of the sex classification model (n=99.095, ‘Abnormal prevalence = 41%) and on a dataset only consisting of the correctly sex-classified patients (n=85.460, ‘abnormal prevalence=39%).

Table: Performance metrics sex classification model

| **AUC** | **Accuracy** | **Precision** | **Recall** | **Loss** |
|---|---|---|---|---|
| 0.94 | 0.86 | 0.85 | 0.87 | 0.33 |

### Table S1

**Table S1:** Characteristics of the included patients and their ECG stratified by sex.

| | Women (n=82,578) | Men (n=82,578) |
|---|---|---|
| **Age (years)** | | |
| 18-40 | 12,563 (15%) | 10,701 (13%) |
| 41-60 | 23,986 (29%) | 24,296 (29%) |
| 61-80 | 36,179 (44%) | 40,338 (49%) |
| > 80 | 9,850 (12%) | 7,243 (9%) |
| **ECG acquisition year** | | |
| ≤ 2000 | 494 (1%) | 537 (1%) |
| 2001-2010 | 32,454 (39%) | 32,347 (39%) |
| 2011-2020 | 35,687 (43%) | 34,789 (42%) |
| > 2020 | 13,943 (17%) | 14,905 (18%) |
| **ECG characteristics** | | |
| Ventricular rate (bpm) | 74 [64, 87] | 72 [62, 85] |
| PR interval (ms) | 152 [138, 170] | 160 [144, 178] |
| QRS duration (ms) | 88 [80, 96] | 98 [88, 106] |
| QT interval (ms) | 384 [360, 412] | 388 [360, 416] |
| QTc interval (ms) | 425 [409, 446] | 422 [406, 445] |
| **Outcome prevalence** | | |
| LBBB | 1,743 (2%) | 1,815 (2%) |
| LQT | 2,365 (3%) | 1,768 (2%) |
| LVH | 3,545 (4%) | 4,911 (6%) |
| Abnormal | 30,731 (37%) | 37,677 (46%) |

*Categorical variables are presented as n (%) and numerical variables as median [Q1, Q3]. LBBB = Left bundle branch block; LVH = Left ventricular hypertrophy; LQT = Long QT syndrome.*

### Table S2

**Table S2A:** Included ECGs in sex-imbalance in representation analysis for a left bundle branch block.

| **Group** | **Sex** | **LBBB** | **Proportion women in training: 1.00** | **Proportion women in training: 0.75** | **Proportion women in training: 0.50** | **Proportion women in training: 0.25** | **Proportion women in training: 0.00** |
|---|---|---|---|---|---|---|---|
| Train | Man | 1 | 0 | 266 | 533 | 800 | 1067 |
| Train | Man | 0 | 0 | 11876 | 23753 | 35630 | 47507 |
| Train | Woman | 1 | 1025 | 768 | 512 | 256 | 0 |
| Train | Woman | 0 | 47549 | 35662 | 23774 | 11887 | 0 |
| Validation | Man | 1 | 0 | 106 | 213 | 320 | 427 |
| Validation | Man | 0 | 0 | 4750 | 9501 | 14252 | 19002 |
| Validation | Woman | 1 | 410 | 307 | 205 | 102 | 0 |
| Validation | Woman | 0 | 19019 | 14264 | 9509 | 4754 | 0 |
| Test | Man | 1 | 320 | 320 | 320 | 320 | 320 |
| Test | Man | 0 | 14252 | 14252 | 14252 | 14252 | 14252 |
| Test | Woman | 1 | 307 | 307 | 307 | 307 | 307 |
| Test | Woman | 0 | 14264 | 14264 | 14264 | 14264 | 14264 |

*The maximum number of patients that could be included in the combined training and test data was determined using Formula 1 (Method S1) and amounted to n = 97,146. LBBB = Left bundle branch block.*

**Table S2B:** Over 5 folds averaged testing metrics for CNN models trained to classify a left bundle branch block.

| **Test metric** | **Sex** | **Proportion women in training: 1.00** | **Proportion women in training: 0.75** | **Proportion women in training: 0.50** | **Proportion women in training: 0.25** | **Proportion women in training: 0.00** |
|---|---|---|---|---|---|---|
| AUC | Men | 0.982 (0.972, 0.991) | 0.986 (0.979, 0.992) | 0.992 (0.991, 0.993) | 0.985 (0.979, 0.992) | 0.991 (0.985, 0.994) |
| AUC | Woman | 0.994 (0.993, 0.996) | 0.995 (0.993, 0.997) | 0.997 (0.996, 0.998) | 0.995 (0.994, 0.997) | 0.993 (0.989, 0.996) |

| | | | | | | |
|---|---|---|---|---|---|---|
| Recall | Men | 0.834 (0.807, 0.86) | 0.868 (0.824, 0.913) | 0.861 (0.838, 0.886) | 0.84 (0.809, 0.867) | 0.848 (0.827, 0.869) |
| Recall | Women | 0.926 (0.911, 0.941) | 0.935 (0.902, 0.957) | 0.939 (0.921, 0.957) | 0.91 (0.878, 0.936) | 0.916 (0.895, 0.936) |
| Precision | Men | 0.762 (0.736, 0.787) | 0.703 (0.65, 0.769) | 0.709 (0.683, 0.735) | 0.735 (0.704, 0.766) | 0.752 (0.728, 0.774) |
| Precision | Women | 0.834 (0.817, 0.854) | 0.828 (0.786, 0.889) | 0.846 (0.812, 0.881) | 0.852 (0.829, 0.885) | 0.853 (0.84, 0.865) |
| smECE | Men | 0.006 (0.006, 0.007) | 0.011 (0.007, 0.015) | 0.01 (0.008, 0.013) | 0.008 (0.007, 0.01) | 0.007 (0.007, 0.008) |
| smECE | Woman | 0.005 (0.004, 0.005) | 0.006 (0.005, 0.008) | 0.005 (0.004, 0.007) | 0.005 (0.005, 0.006) | 0.004 (0.004, 0.004) |

*Models were trained with different proportions of women and men included in the training data. For each proportion and fold the training data included around 48,574 ECGs, validation included around 19,429 ECGs, and test data included around 29,143 ECGs. LBBB = Left bundle branch block.*

### Table S3

**Table S3A:** Included ECGs in sex-imbalance in representation analysis for Long QT Syndrome.

| Group | Sex | LQT | Proportion women in training: 1.00 | Proportion women in training: 0.75 | Proportion women in training: 0.50 | Proportion women in training: 0.25 | Proportion women in training: 0.00 |
|---|---|---|---|---|---|---|---|
| Train | Man | 1 | 0 | 259 | 519 | 779 | 1039 |
| Train | Man | 0 | 0 | 11883 | 23767 | 35651 | 47535 |
| Train | Woman | 1 | 1391 | 1043 | 695 | 347 | 0 |
| Train | Woman | 0 | 47183 | 35387 | 23591 | 11795 | 0 |
| Validation | Man | 1 | 0 | 103 | 207 | 311 | 415 |
| Validation | Man | 0 | 0 | 4753 | 9507 | 14260 | 19014 |
| Validation | Woman | 1 | 556 | 417 | 278 | 139 | 0 |
| Validation | Woman | 0 | 18873 | 14155 | 9436 | 4718 | 0 |
| Test | Man | 1 | 311 | 311 | 311 | 311 | 311 |
| Test | Man | 0 | 14260 | 14260 | 14260 | 14260 | 14260 |
| Test | Woman | 1 | 417 | 417 | 417 | 417 | 417 |
| Test | Woman | 0 | 14155 | 14155 | 14155 | 14155 | 14155 |

*The maximum number of patients that could be included in the combined training and test data was determined using Formula 1 (Method S1) and amounted to n = 97,146. LQT = Long QT Syndrome.*

**Table S3B:** Over 5 folds averaged testing metrics for CNN models trained to classify Long QT Syndrome.

| Test metric | Sex | Proportion women in training: 1.00 | Proportion women in training: 0.75 | Proportion women in training: 0.50 | Proportion women in training: 0.25 | Proportion women in training: 0.00 |
|---|---|---|---|---|---|---|
| AUC | Men | 0.892 (0.885, 0.899) | 0.882 (0.861, 0.898) | 0.893 (0.884, 0.9) | 0.888 (0.869, 0.899) | 0.891 (0.882, 0.896) |
| AUC | Women | 0.898 (0.89, 0.906) | 0.894 (0.878, 0.903) | 0.898 (0.893, 0.902) | 0.894 (0.875, 0.907) | 0.876 (0.863, 0.892) |

| | | | | | | |
|---|---|---|---|---|---|---|
| Recall | Men | 0.215 (0.158, 0.271) | 0.196 (0.149, 0.251) | 0.175 (0.113, 0.221) | 0.164 (0.105, 0.224) | 0.134 (0.084, 0.185) |
| Recall | Women | 0.215 (0.138, 0.289) | 0.219 (0.159, 0.262) | 0.218 (0.157, 0.26) | 0.217 (0.136, 0.302) | 0.163 (0.091, 0.235) |
| Precision | Men | 0.262 (0.243, 0.282) | 0.295 (0.26, 0.333) | 0.293 (0.285, 0.305) | 0.309 (0.289, 0.338) | 0.317 (0.281, 0.35) |
| Precision | Women | 0.36 (0.316, 0.396) | 0.399 (0.349, 0.443) | 0.435 (0.394, 0.498) | 0.414 (0.36, 0.472) | 0.426 (0.398, 0.45) |
| smECE | Men | 0.023 (0.017, 0.028) | 0.02 (0.016, 0.024) | 0.022 (0.018, 0.025) | 0.016 (0.014, 0.018) | 0.018 (0.012, 0.024) |
| smECE | Woman | 0.019 (0.012, 0.026) | 0.018 (0.011, 0.024) | 0.022 (0.019, 0.025) | 0.017 (0.013, 0.021) | 0.02 (0.012, 0.028) |

*Models were trained with different proportions of women and men included in the training data. For each proportion and fold the training data included around 48,574 ECGs, validation included around 19,429 ECGs, and test data included around 29,143 ECGs. LQT = Long QT Syndrome.*

### Table S4

**Table S4A:** Included ECGs in sex-imbalance in representation analysis for left ventricular hypertrophy.

| Group | Sex | LVH | Proportion women in training: 1.00 | Proportion women in training: 0.75 | Proportion women in training: 0.50 | Proportion women in training: 0.25 | Proportion women in training: 0.00 |
|---|---|---|---|---|---|---|---|
| Train | Man | 1 | 0 | 722 | 1444 | 2166 | 2888 |
| Train | Man | 0 | 0 | 11421 | 22843 | 34264 | 45686 |
| Train | Woman | 1 | 2085 | 1563 | 1042 | 521 | 0 |
| Train | Woman | 0 | 46489 | 34867 | 23244 | 11622 | 0 |
| Validation | Man | 1 | 0 | 288 | 577 | 866 | 1155 |
| Validation | Man | 0 | 0 | 4568 | 9137 | 13705 | 18274 |
| Validation | Woman | 1 | 834 | 625 | 417 | 208 | 0 |
| Validation | Woman | 0 | 18595 | 13946 | 9297 | 4648 | 0 |
| Test | Man | 1 | 866 | 866 | 866 | 866 | 866 |
| Test | Man | 0 | 13705 | 13705 | 13705 | 13705 | 13705 |
| Test | Woman | 1 | 625 | 625 | 625 | 625 | 625 |
| Test | Woman | 0 | 13946 | 13946 | 13946 | 13946 | 13946 |

*The maximum number of patients that could be included in the combined training and test data was determined using Formula 1 (Method S1) and amounted to n = 97,146. LVH = Left ventricular hypertrophy.*

**Table S4B:** Over 5 folds averaged testing metrics for CNN models trained to classify left ventricular hypertrophy.

| Test metric | Sex | Proportion women in training: 1.00 | Proportion women in training: 0.75 | Proportion women in training: 0.50 | Proportion women in training: 0.25 | Proportion women in training: 0.00 |
|---|---|---|---|---|---|---|
| AUC | Men | 0.948 (0.946,0.951) | 0.948 (0.944, 0.951) | 0.954 (0.952, 0.957) | 0.955 (0.951, 0.96) | 0.954 (0.95, 0.957) |
| AUC | Women | 0.968 (0.967 , 0.969) | 0.964 (0.961, 0.966) | 0.966 (0.961, 0.97) | 0.967 (0.966, 0.968) | 0.967 (0.964 , 0.969) |

| | | | | | | |
|---|---|---|---|---|---|---|
| Recall | Men | 0.544 (0.506, 0.574) | 0.485 (0.435, 0.531) | 0.464 (0.438, 0.482) | 0.48 (0.445, 0.502) | 0.501 (0.437 , 0.546) |
| Recall | Women | 0.55 (0.533, 0.57) | 0.518 (0.466, 0.57) | 0.483 (0.427, 0.524) | 0.457 (0.432, 0.49) | 0.46 (0.422, 0.501) |
| Precision | Men | 0.602 (0.577, 0.627) | 0.644 (0.615, 0.668) | 0.687 (0.656, 0.715) | 0.72 (0.7, 0.739) | 0.676 (0.649 , 0.706) |
| Precision | Women | 0.683 (0.653, 0.709) | 0.664 (0.624, 0.701) | 0.732 (0.708, 0.76) | 0.746 (0.729, 0.762) | 0.69 (0.658, 0.731) |
| smECE | Men | 0.024 (0.017, 0.03) | 0.014 (0.011, 0.016) | 0.012 (0.011, 0.013) | 0.01 (0.009, 0.012) | 0.012 (0.011 ,0.013) |
| smECE | Women | 0.012 (0.009, 0.015) | 0.01 (0.009, 0.012) | 0.009 (0.008, 0.01) | 0.01 (0.009, 0.011) | 0.011 (0.009 ,0.012) |

*Models were trained with different proportions of women and men included in the training data. For each proportion and fold the training data included around 48,574 ECGs, validation included around 19,429 ECGs, and test data included around 29,143 ECGs. LVH = Left ventricular hypertrophy.*

### Table S5

**Table S5A:** Included ECGs in sex-imbalance in representation analysis for the outcome 'abnormal'.

| Group | Sex | abnormal | Proportion women in training: 1.00 | Proportion women in training: 0.75 | Proportion women in training: 0.50 | Proportion women in training: 0.25 | Proportion women in training: 0.00 |
|---|---|---|---|---|---|---|---|
| Train | Man | 1 | 0 | 5540 | 11081 | 16622 | 22162 |
| Train | Man | 0 | 0 | 6603 | 13206 | 19809 | 26412 |
| Train | Woman | 1 | 18076 | 13557 | 9038 | 4519 | 0 |
| Train | Woman | 0 | 30498 | 22873 | 15249 | 7624 | 0 |
| Validation | Man | 1 | 0 | 2216 | 4432 | 6648 | 8865 |
| Validation | Man | 0 | 0 | 2641 | 5282 | 7923 | 10564 |
| Validation | Woman | 1 | 7230 | 5423 | 3615 | 1807 | 0 |
| Validation | Woman | 0 | 12199 | 9149 | 6099 | 3049 | 0 |
| Test | Man | 1 | 5423 | 5423 | 5423 | 5423 | 5423 |
| Test | Man | 0 | 9149 | 9149 | 9149 | 9149 | 9149 |
| Test | Woman | 1 | 6648 | 6648 | 6648 | 6648 | 6648 |
| Test | Woman | 0 | 7923 | 7923 | 7923 | 7923 | 7923 |

*The maximum number of patients that could be included in the combined training and test data was determined using Formula 1 (Method S1) and amounted to n = 97,146.*

**Table S5B:** Over 5 folds averaged testing metrics for CNN models trained to classify 'abnormal'.

| Test metric | Sex | Proportion women in training: 1.00 | Proportion women in training: 0.75 | Proportion women in training: 0.50 | Proportion women in training: 0.25 | Proportion women in training: 0.00 |
|---|---|---|---|---|---|---|
| AUC | Men | 0.882 (0.874 , 0.887) | 0.888 (0.885, 0.891) | 0.892 (0.889, 0.894) | 0.893 (0.889, 0.9) | 0.893 (0.888, 0.897) |
| AUC | Women | 0.907 (0.9, 0.913) | 0.906 (0.902, 0.91) | 0.904 (0.901, 0.907) | 0.905 (0.903, 0.906) | 0.898 (0.896, 0.9) |

| | | | | | | |
|---|---|---|---|---|---|---|
| Recall | Men | 0.677 (0.632, 0.712) | 0.734 (0.724, 0.746) | 0.711 (0.695, 0.735) | 0.706 (0.695, 0.717) | 0.701 (0.687, 0.715) |
| Recall | Women | 0.674 (0.633, 0.705) | 0.696 (0.68, 0.713) | 0.666 (0.648, 0.69) | 0.657 (0.655, 0.66) | 0.651 (0.64, 0.663) |
| Precision | Men | 0.854 (0.843, 0.863) | 0.838 (0.827, 0.848) | 0.852 (0.849, 0.856) | 0.856 (0.848, 0.864) | 0.856 (0.85, 0.862) |
| Precision | Women | 0.85 (0.839, 0.861) | 0.83 (0.823, 0.838) | 0.842 (0.831, 0.852) | 0.848 (0.844, 0.852) | 0.838 (0.829, 0.847) |
| smECE | Men | 0.075 (0.061, 0.091) | 0.049 (0.037, 0.06) | 0.059 (0.05, 0.064) | 0.061 (0.055, 0.066) | 0.063 (0.056, 0.07) |
| smECE | Women | 0.042 (0.035, 0.05) | 0.037 (0.029, 0.044) | 0.045 (0.035, 0.053) | 0.048 (0.044, 0.051) | 0.044 (0.038, 0.049) |

*Models were trained with different proportions of women and men included in the training data. For each proportion and fold the training data included around 48,574 ECGs, validation included around 19,429 ECGs, and test data included around 29,143 ECGs.*

**Table S6**

**Table S6A:** Included ECGs in sex-imbalance in 'abnormal' prevalence analysis.

| Group | Sex | abnormal | Prevalence women: 90% men: 10% | Prevalence women: 70% men: 30% | Prevalence women: 50% men: 50% | Prevalence women: 30% men: 70% | Prevalence women: 10% men: 90% |
|---|---|---|---|---|---|---|---|
| Train | Man | 1 | 1707 | 5121 | 8536 | 11950 | 15365 |
| Train | Man | 0 | 15365 | 11950 | 8536 | 5121 | 1707 |
| Train | Woman | 1 | 15365 | 11950 | 8536 | 5121 | 1707 |
| Train | Woman | 0 | 1707 | 5121 | 8536 | 11950 | 15365 |
| Validation | Man | 1 | 682 | 2048 | 3414 | 4780 | 6146 |
| Validation | Man | 0 | 6146 | 4780 | 3414 | 2048 | 682 |
| Validation | Woman | 1 | 6146 | 4780 | 3414 | 2048 | 682 |
| Validation | Woman | 0 | 682 | 2048 | 3414 | 4780 | 6146 |
| Test | Man | 1 | 1024 | 3073 | 5121 | 7170 | 9219 |
| Test | Man | 0 | 9219 | 7170 | 5121 | 3073 | 1024 |
| Test | Woman | 1 | 9219 | 7170 | 5121 | 3073 | 1024 |
| Test | Woman | 0 | 1024 | 3073 | 5121 | 7170 | 9219 |

*The maximum number of patients that could be included in the combined training and test data was determined using Formula 1 (Method S1) and amounted to n = 68,286.*

**Table S6B:** Over 5 folds averaged testing metrics for CNN models trained to classify 'abnormal'.

| Test metric | Sex | Prevalence women:90 % men: 10% | Prevalence women:70 % men: 30% | Prevalence women: 50% men: 50% | Prevalence women:30 % men: 70% | Prevalence women:10 % men: 90% |
|---|---|---|---|---|---|---|
| AUC | Men | 0.835 (0.821, 0.848) | 0.872 (0.864, 0.879) | 0.889 (0.886, 0.892) | 0.884 (0.881, 0.888) | 0.848 (0.841, 0.855) |
| AUC | Women | 0.856 (0.848, 0.864) | 0.887 (0.882, 0.892) | 0.901 (0.897, 0.905) | 0.896 (0.894, 0.898) | 0.869 (0.863, 0.875) |
| Recall | Men | 0.529 (0.479, 0.581) | 0.616 (0.575, 0.647) | 0.729 (0.711, 0.752) | 0.818 (0.798, 0.836) | 0.879 (0.868, 0.891) |
| Recall | Women | 0.818 (0.795, 0.849) | 0.728 (0.682, 0.77) | 0.72 (0.708, 0.732) | 0.671 (0.651, 0.682) | 0.591 (0.56, 0.623) |
| Precision | Men | 0.422 (0.403, 0.441) | 0.772 (0.763, 0.781) | 0.868 (0.859, 0.875) | 0.909 (0.904, 0.918) | 0.954 (0.951, 0.958) |

| | | | | | | |
|---|---|---|---|---|---|---|
| Precision | Women | 0.965 (0.958, 0.971) | 0.935 (0.93, 0.94) | 0.881 (0.872, 0.888) | 0.779 (0.764, 0.794) | 0.487 (0.472, 0.501) |
| smECE | Men | 0.071 (0.058, 0.091) | 0.044 (0.034, 0.055) | 0.061 (0.051, 0.067) | 0.074 (0.063, 0.09) | 0.107 (0.094, 0.116) |
| smECE | Women | 0.178 (0.144, 0.201) | 0.15 (0.124, 0.177) | 0.063 (0.055, 0.069) | 0.028 (0.027, 0.029) | 0.074 (0.064, 0.085) |

*Models were trained with different proportions of women and men included in the training data. For each proportion and fold the training data included around 34,144 ECGs, validation included around 13,656 ECGs, and test data included around 20,486 ECGs.*

### Table S7

**Table S7A:** Included ECGs in sex-imbalance in 'abnormal' representation and prevalence analysis (analysis 3 scenarios 1-5: 90% in women; 50% in men).

| Group | Sex | Abnormal | Proportion women in training: 1.00 | Proportion women in training: 0.75 | Proportion women in training: 0.50 | Proportion women in training: 0.25 | Proportion women in training: 0.00 |
|---|---|---|---|---|---|---|---|
| Train | Man | 1 | 0 | 2510 | 5021 | 7532 | 10042 |
| Train | Man | 0 | 0 | 2510 | 5021 | 7532 | 10042 |
| Train | Woman | 1 | 18076 | 13557 | 9038 | 4519 | 0 |
| Train | Woman | 0 | 2008 | 1506 | 1004 | 502 | 0 |
| Validation | Man | 1 | 0 | 1004 | 2008 | 3012 | 4017 |
| Validation | Man | 0 | 0 | 1004 | 2008 | 3012 | 4017 |
| Validation | Woman | 1 | 7230 | 5423 | 3615 | 1807 | 0 |
| Validation | Woman | 0 | 803 | 602 | 401 | 200 | 0 |
| Test | Man | 1 | 3012 | 3012 | 3012 | 3012 | 3012 |
| Test | Man | 0 | 3012 | 3012 | 3012 | 3012 | 3012 |
| Test | Woman | 1 | 5423 | 5423 | 5423 | 5423 | 5423 |
| Test | Woman | 0 | 602 | 602 | 602 | 602 | 602 |

*The maximum number of patients that could be included in the combined training and test data was determined using Formula 1 (Method S1) and amounted to n = 40,167.*

**Table S7B:** Over 5 folds averaged testing metrics for CNN models trained to classify 'abnormal' (analysis 3 scenarios 1-5: 90% in women; 50% in men).

| Test metric | Sex | Proportion women in training: 1.00 | Proportion women in training: 0.75 | Proportion women in training: 0.50 | Proportion women in training: 0.25 | Proportion women in training: 0.00 |
|---|---|---|---|---|---|---|
| AUC | Men | 0.865 (0.86, 0.871) | 0.852 (0.844, 0.86) | 0.868 (0.862, 0.873) | 0.866 (0.864, 0.868) | 0.879 (0.872, 0.886) |
| AUC | Women | 0.904 (0.9, 0.909) | 0.881 (0.867, 0.894) | 0.89 (0.882, 0.895) | 0.88 (0.877, 0.883) | 0.897 (0.894, 0.9) |

| | | | | | | |
|---|---|---|---|---|---|---|
| Recall | Men | 0.876 (0.86, 0.892) | 0.734 (0.71, 0.758) | 0.707 (0.683, 0.736) | 0.682 (0.662, 0.697) | 0.685 (0.653, 0.711) |
| Recall | Women | 0.906 (0.898, 0.915) | 0.87 (0.851, 0.884) | 0.828 (0.807, 0.85) | 0.795 (0.778, 0.811) | 0.65 (0.632, 0.669) |
| Precision | Men | 0.725 (0.708, 0.744) | 0.815 (0.798, 0.834) | 0.852 (0.839, 0.87) | 0.858 (0.848, 0.865) | 0.865 (0.855, 0.873) |
| Precision | Women | 0.962 (0.959, 0.964) | 0.962 (0.955, 0.968) | 0.972 (0.967, 0.978) | 0.973 (0.97, 0.976) | 0.988 (0.986, 0.99) |
| smECE | Men | 0.123 (0.106, 0.146) | 0.051 (0.039, 0.065) | 0.076 (0.06, 0.091) | 0.072 (0.062, 0.08) | 0.079 (0.072, 0.088) |
| smECE | Women | 0.093 (0.08, 0.103) | 0.142 (0.132, 0.152) | 0.177 (0.162, 0.196) | 0.21 (0.196, 0.228) | 0.291 (0.283, 0.298) |

*Models were trained with different proportions of women and men included in the training data. For each proportion and fold the training data included around 20,084 ECGs, validation included around 8,034 ECGs, and test data included around 12,049 ECGs.*

### Table S8

**Table S8A:** Included ECGs in sex-imbalance in 'abnormal' representation and prevalence analysis (analysis 3 scenario 6-10: 50% in women; 10% in men).

| Group | Sex | abnormal | Proportion women in training: 1.00 | Proportion women in training: 0.75 | Proportion women in training: 0.50 | Proportion women in training: 0.25 | Proportion women in training: 0.00 |
|---|---|---|---|---|---|---|---|
| Train | Man | 1 | 0 | 502 | 1004 | 1506 | 2008 |
| Train | Man | 0 | 0 | 4519 | 9038 | 13557 | 18076 |
| Train | Woman | 1 | 10042 | 7532 | 5021 | 2510 | 0 |
| Train | Woman | 0 | 10042 | 7532 | 5021 | 2510 | 0 |
| Validation | Man | 1 | 0 | 200 | 401 | 602 | 803 |
| Validation | Man | 0 | 0 | 1807 | 3615 | 5423 | 7230 |
| Validation | Woman | 1 | 4017 | 3012 | 2008 | 1004 | 0 |
| Validation | Woman | 0 | 4017 | 3012 | 2008 | 1004 | 0 |
| Test | Man | 1 | 602 | 602 | 602 | 602 | 602 |
| Test | Man | 0 | 5423 | 5423 | 5423 | 5423 | 5423 |
| Test | Woman | 1 | 3012 | 3012 | 3012 | 3012 | 3012 |
| Test | Woman | 0 | 3012 | 3012 | 3012 | 3012 | 3012 |

*The maximum number of patients that could be included in the combined training and test data was determined using Formula 1 (Method S1) and amounted to n = 40,167.*

**Table S8B:** Over 5 folds averaged testing metrics for CNN models trained to classify 'abnormal' (analysis 3 scenario 6-10: 50% in women 10% in men.

| Test metric | Sex | Proportion women in training: 1.00 | Proportion women in training: 0.75 | Proportion women in training: 0.50 | Proportion women in training: 0.25 | Proportion women in training: 0.00 |
|---|---|---|---|---|---|---|
| AUC | Men | 0.879 (0.874, 0.884) | 0.861 (0.852, 0.867) | 0.854 (0.845, 0.862) | 0.861 (0.855, 0.866) | 0.85 (0.846, 0.856) |
| AUC | Women | 0.893 (0.89, 0.896) | 0.888 (0.884, 0.893) | 0.872 (0.863, 0.879) | 0.867 (0.858, 0.874) | 0.869 (0.86, 0.878) |
| Recall | Men | 0.713 (0.685, 0.745) | 0.561 (0.547, 0.579) | 0.485 (0.471, 0.501) | 0.409 (0.392, 0.424) | 0.283 (0.263, 0.308) |

| | | | | | | |
|---|---|---|---|---|---|---|
| Recall | Women | 0.724 (0.699, 0.742) | 0.658 (0.634, 0.683) | 0.614 (0.594, 0.644) | 0.518 (0.482, 0.555) | 0.264 (0.238, 0.287) |
| Precision | Men | 0.399 (0.366, 0.433) | 0.49 (0.463, 0.505) | 0.519 (0.506, 0.537) | 0.569 (0.54, 0.588) | 0.601 (0.576, 0.627) |
| Precision | Women | 0.867 (0.858, 0.876) | 0.881 (0.87, 0.893) | 0.881 (0.869, 0.894) | 0.906 (0.897, 0.915) | 0.948 (0.93, 0.962) |
| smECE | Men | 0.153 (0.134, 0.172) | 0.067 (0.059, 0.072) | 0.042 (0.039, 0.044) | 0.024 (0.021, 0.027) | 0.018 (0.016, 0.019) |
| smECE | Women | 0.068 (0.056, 0.079) | 0.097 (0.082, 0.11) | 0.117 (0.107, 0.124) | 0.169 (0.152, 0.183) | 0.279 (0.272, 0.285) |

*Models were trained with different proportions of women and men included in the training data. For each proportion and fold the training data included around 20,084 ECGs, validation included around 8,034 ECGs, and test data included around 12,049 ECGs.*

### Table S9

**Table S9A:** Included ECGs in sex-imbalance in 'abnormal' representation and prevalence analysis (analysis 3 scenarios 11-15: 50% in women; 90% in men).

| Group | Sex | abnormal | Proportion women in training: 1.00 | Proportion women in training: 0.75 | Proportion women in training: 0.50 | Proportion women in training: 0.25 | Proportion women in training: 0.00 |
|---|---|---|---|---|---|---|---|
| Train | Man | 1 | 0 | 4519 | 9038 | 13557 | 18076 |
| Train | Man | 0 | 0 | 502 | 1004 | 1506 | 2008 |
| Train | Woman | 1 | 10042 | 7532 | 5021 | 2510 | 0 |
| Train | Woman | 0 | 10042 | 7532 | 5021 | 2510 | 0 |
| Validation | Man | 1 | 0 | 1807 | 3615 | 5423 | 7230 |
| Validation | Man | 0 | 0 | 200 | 401 | 602 | 803 |
| Validation | Woman | 1 | 4017 | 3012 | 2008 | 1004 | 0 |
| Validation | Woman | 0 | 4017 | 3012 | 2008 | 1004 | 0 |
| Test | Man | 1 | 5423 | 5423 | 5423 | 5423 | 5423 |
| Test | Man | 0 | 602 | 602 | 602 | 602 | 602 |
| Test | Woman | 1 | 3012 | 3012 | 3012 | 3012 | 3012 |
| Test | Woman | 0 | 3012 | 3012 | 3012 | 3012 | 3012 |

*The maximum number of patients that could be included in the combined training and test data was determined using Formula 1 (Method S1) and amounted to n = 40,167.*

**Table S9B:** Over 5 folds averaged testing metrics for CNN models trained to classify 'abnormal' (analysis 3 scenarios 11-15 : 50% in women; 90% in men).

| Test metric | Sex | Proportion women in training: 1.00 | Proportion women in training: 0.75 | Proportion women in training: 0.50 | Proportion women in training: 0.25 | Proportion women in training: 0.00 |
|---|---|---|---|---|---|---|
| AUC | Men | 0.878 (0.865, 0.886) | 0.867 (0.862, 0.871) | 0.875 (0.863, 0.885) | 0.867 (0.856, 0.879) | 0.867 (0.855, 0.88) |
| AUC | Women | 0.896 (0.893, 0.9) | 0.895 (0.892, 0.897) | 0.885 (0.879, 0.89) | 0.885 (0.88, 0.889) | 0.87 (0.861, 0.877) |

| | | | | | | |
|---|---|---|---|---|---|---|
| Recall | Men | 0.731 (0.696, 0.767) | 0.857 (0.834, 0.877) | 0.87 (0.854, 0.886) | 0.89 (0.877, 0.906) | 0.909 (0.896, 0.923) |
| Recall | Women | 0.708 (0.686, 0.74) | 0.741 (0.721, 0.763) | 0.743 (0.717, 0.778) | 0.759 (0.744, 0.775) | 0.928 (0.896, 0.957) |
| Precision | Men | 0.982 (0.98, 0.983) | 0.965 (0.963, 0.966) | 0.967 (0.964, 0.969) | 0.961 (0.958, 0.963) | 0.955 (0.945, 0.963) |
| Precision | Women | 0.872 (0.86, 0.883) | 0.855 (0.843, 0.867) | 0.84 (0.829, 0.85) | 0.83 (0.818, 0.838) | 0.651 (0.594, 0.7) |
| smECE | Man | 0.234 (0.21, 0.257) | 0.129 (0.107, 0.15) | 0.119 (0.101, 0.137) | 0.101 (0.094, 0.109) | 0.103 (0.097, 0.11) |
| smECE | Woman | 0.067 (0.053, 0.078) | 0.053 (0.047, 0.058) | 0.053 (0.045, 0.06) | 0.043 (0.034, 0.051) | 0.162 (0.129, 0.199) |

*Models were trained with different proportions of women and men included in the training data. For each proportion and fold the training data included around 20,084 ECGs, validation included around 8,034 ECGs, and test data included around 12,049 ECGs.*

### Table S10

**Table S10A:** Included ECGs in sex-imbalance in 'abnormal' representation and prevalence analysis (analysis 3 scenarios 16-20: 10% in women; 50% in men).

| Group | Sex | Abnormal | Proportion women in training: 1.00 | Proportion women in training: 0.75 | Proportion women in training: 0.50 | Proportion women in training: 0.25 | Proportion women in training: 0.00 |
|---|---|---|---|---|---|---|---|
| Train | Man | 1 | 0 | 2510 | 5021 | 7532 | 10042 |
| Train | Man | 0 | 0 | 2510 | 5021 | 7532 | 10042 |
| Train | Woman | 1 | 2008 | 1506 | 1004 | 502 | 0 |
| Train | Woman | 0 | 18076 | 13557 | 9038 | 4519 | 0 |
| Validation | Man | 1 | 0 | 1004 | 2008 | 3012 | 4017 |
| Validation | Man | 0 | 0 | 1004 | 2008 | 3012 | 4017 |
| Validation | Woman | 1 | 803 | 602 | 401 | 200 | 0 |
| Validation | Woman | 0 | 7230 | 5423 | 3615 | 1807 | 0 |
| Test | Man | 1 | 3012 | 3012 | 3012 | 3012 | 3012 |
| Test | Man | 0 | 3012 | 3012 | 3012 | 3012 | 3012 |
| Test | Woman | 1 | 602 | 602 | 602 | 602 | 602 |
| Test | Woman | 0 | 5423 | 5423 | 5423 | 5423 | 5423 |

*The maximum number of patients that could be included in the combined training and test data was determined using Formula 1 (Method S1) and amounted to n = 40,167.*

**Table S10B:** Over 5 folds averaged testing metrics for CNN models trained to classify 'abnormal' (analysis 3 scenarios 16-20: 10% in women; 50% in men).

| Test metric | Sex | Proportion women in training: 1.00 | Proportion women in training: 0.75 | Proportion women in training: 0.50 | Proportion women in training: 0.25 | Proportion women in training: 0.00 |
|---|---|---|---|---|---|---|
| AUC | Men | 0.851 (0.843, 0.858) | 0.862 (0.857, 0.867) | 0.872 (0.864, 0.877) | 0.874 (0.87, 0.878) | 0.877 (0.873, 0.884) |
| AUC | Women | 0.881 (0.871, 0.894) | 0.878 (0.872, 0.885) | 0.879 (0.871, 0.887) | 0.888 (0.875, 0.9) | 0.888 (0.881, 0.894) |

| | | | | | | |
|---|---|---|---|---|---|---|
| Recall | Men | 0.336 (0.312, 0.354) | 0.609 (0.585, 0.634) | 0.67 (0.649, 0.681) | 0.673 (0.648, 0.7) | 0.676 (0.65, 0.703) |
| Recall | Women | 0.327 (0.298, 0.354) | 0.454 (0.428, 0.477) | 0.486 (0.466, 0.508) | 0.572 (0.526, 0.611) | 0.641 (0.613, 0.667) |
| Precision | Men | 0.923 (0.912, 0.934) | 0.877 (0.867, 0.885) | 0.867 (0.857, 0.881) | 0.866 (0.862, 0.874) | 0.872 (0.865, 0.876) |
| Precision | Women | 0.687 (0.661, 0.713) | 0.603 (0.579, 0.631) | 0.595 (0.574, 0.61) | 0.561 (0.539, 0.578) | 0.484 (0.47, 0.497) |
| smECE | Men | 0.259 (0.253, 0.266) | 0.118 (0.103, 0.132) | 0.089 (0.078, 0.1) | 0.085 (0.072, 0.093) | 0.08 (0.069, 0.095) |
| smECE | Women | 0.02 (0.017, 0.023) | 0.024 (0.021, 0.027) | 0.036 (0.026, 0.045) | 0.065 (0.057, 0.075) | 0.134 (0.125, 0.141) |

*Models were trained with different proportions of women and men included in the training data. For each proportion and fold the training data included around 20,084 ECGs, validation included around 8,034 ECGs, and test data included around 12,049 ECGs.*

### Table S11

**Table S11A:** Included ECGs in sex-imbalance in ‘abnormal’ misclassification analysis.

| Group | Sex | abnormal shuffled | Outcomes in training data shuffled | | | | | | |
|---|---|---|---|---|---|---|---|---|---|
| | | | 100% women | 50% women | 10% women | 0% | 10% men | 50% men | 100% men |
| Train | Man | 1 | 30730 | 15365 | 3073 | 0 | 0 | 0 | 0 |
| Train | Man | 0 | 0 | 15365 | 27657 | 30730 | 30730 | 30730 | 30730 |
| Train | Woman | 1 | 0 | 0 | 0 | 0 | 3037 | 15365 | 30730 |
| Train | Woman | 0 | 30730 | 30730 | 30730 | 30730 | 27657 | 15365 | 0 |
| Validation | Man | 1 | 12292 | 6146 | 1229 | 0 | 0 | 0 | 0 |
| Validation | Man | 0 | 0 | 6146 | 11063 | 12292 | 12292 | 12292 | 12292 |
| Validation | Woman | 1 | 0 | 0 | 0 | 0 | 1229 | 6146 | 12292 |
| Validation | Woman | 0 | 12282 | 12292 | 12292 | 12292 | 11063 | 6146 | 0 |
| Test | Man | 0 | 18438 | 18438 | 18438 | 18438 | 18438 | 18438 | 18438 |
| Test | Woman | 0 | 18438 | 18438 | 18438 | 18438 | 18438 | 18438 | 18438 |

*The maximum number of patients that could be included in the combined training and test data was determined using Formula 1 (Method S1) and amounted to n = 122,920.* The proportion of women and men was equal in training, testing and validation data. The prevalence of
the predicted outcome ‘abnormal’ was 50% for both sexes.

**Table S11B:** Over 5 folds averaged testing metrics for CNN models trained to classify ‘abnormal’.

| Group | Sex | Outcomes in training data shuffled | | | | | | |
|---|---|---|---|---|---|---|---|---|
| | | 100% of women | 50% of women | 10% of women | 0% | 10% of men | 50% of men | 100% of men |
| AUC | Men | 0.858 (0.854, 0.864) | 0.879 (0.876, 0.882) | 0.891 (0.888, 0.895) | 0.891 (0.889, 0.893) | 0.89 (0.887, 0.895) | 0.869 (0.866, 0.873) | 0.797 (0.791, 0.805) |
| AUC | Woman | 0.815 (0.8, 0.823) | 0.887 (0.879, 0.892) | 0.905 (0.903, 0.907) | 0.906 (0.903, 0.909) | 0.906 (0.905, 0.907) | 0.895 (0.892, 0.9) | 0.873 (0.87, 0.878) |
| Recall | Men | 0.701 (0.687, 0.716) | 0.721 (0.693, 0.749) | 0.732 (0.719, 0.743) | 0.738 (0.712, 0.77) | 0.743 (0.714, 0.762) | 0.695 (0.679, 0.71) | 0.654 (0.632, 0.675) |
| Recall | Women | 0.634 (0.598, 0.669) | 0.694 (0.664, 0.724) | 0.709 (0.695, 0.723) | 0.714 (0.683, 0.743) | 0.729 (0.705, 0.753) | 0.689 (0.682, 0.696) | 0.686 (0.68, 0.693) |
| Precision | Men | 0.84 (0.83, 0.85) | 0.858 (0.848, 0.869) | 0.87 (0.867, 0.873) | 0.864 (0.849, 0.876) | 0.862 (0.85, 0.874) | 0.855 (0.85, 0.859) | 0.766 (0.748, 0.79) |

| | | | | | | | | |
|---|---|---|---|---|---|---|---|---|
| Precision | Women | 0.81 (0.791, 0.828) | 0.874 (0.864, 0.886) | 0.891 (0.887, 0.897) | 0.89 (0.878, 0.898) | 0.884 (0.877, 0.89) | 0.885 (0.88, 0.89) | 0.858 (0.85, 0.867) |
| smECE | Men | 0.079 (0.074, 0.085) | 0.064 (0.058, 0.074) | 0.058 (0.052, 0.062) | 0.06 (0.049, 0.072) | 0.05 (0.041, 0.064) | 0.08 (0.075, 0.084) | 0.081 (0.076, 0.086) |
| smECE | Women | 0.091 (0.086, 0.096) | 0.095 (0.089, 0.103) | 0.068 (0.062, 0.075) | 0.072 (0.058, 0.085) | 0.063 (0.052, 0.072) | 0.085 (0.082, 0.09) | 0.096 (0.093, 0.1) |

*Models were trained with different proportions of women and men included in the training data. For each proportion and fold the training data included around 61,460 ECGs, validation included around 24,584 ECGs, and test data included around 36,876 ECGs.*

### Figure S1

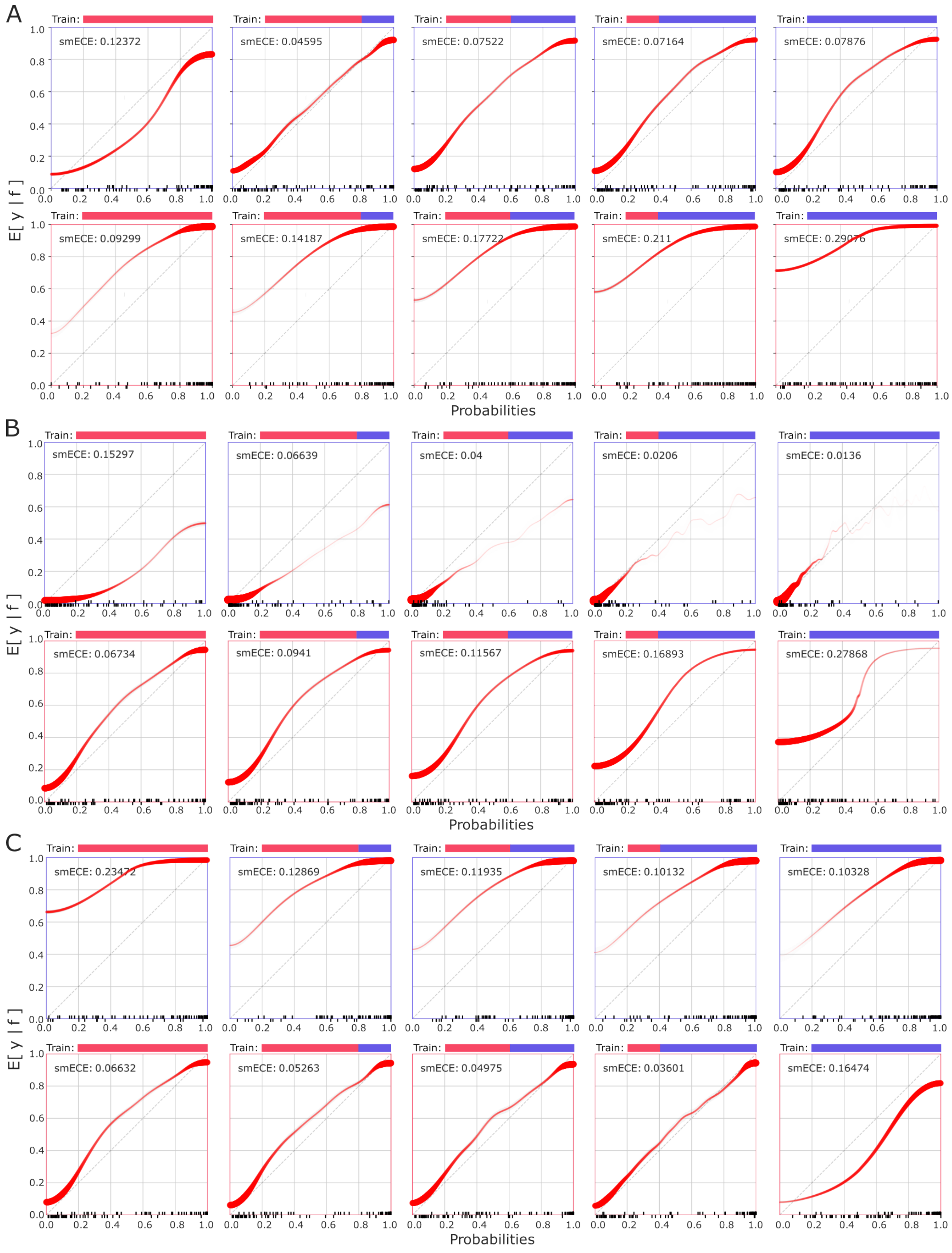
A
Train:
smECE: 0.12372
smECE: 0.04595
smECE: 0.07522
smECE: 0.07164
smECE: 0.07876
smECE: 0.09299
smECE: 0.14187
smECE: 0.17722
smECE: 0.211
smECE: 0.29076
E[ y | f ]
Probabilities
B
smECE: 0.15297
smECE: 0.06639
smECE: 0.04
smECE: 0.0206
smECE: 0.0136
smECE: 0.06734
smECE: 0.0941
smECE: 0.11567
smECE: 0.16893
smECE: 0.27868
C
smECE: 0.23472
smECE: 0.12869
smECE: 0.11935
smECE: 0.10132
smECE: 0.10328
smECE: 0.06632
smECE: 0.05263
smECE: 0.04975
smECE: 0.03601
smECE: 0.16474

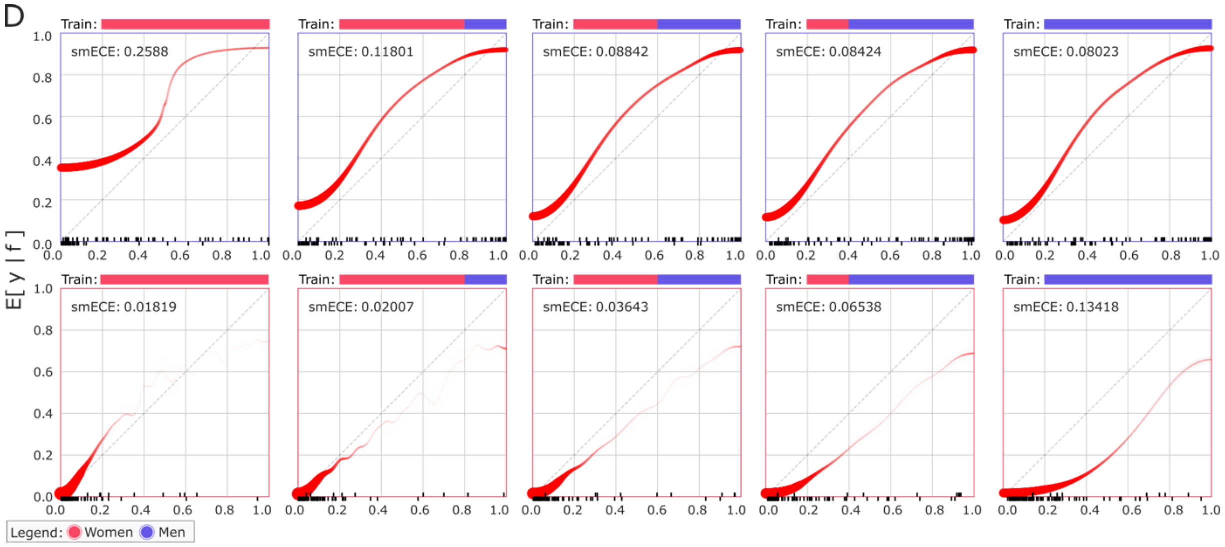


***Figure S1: Calibration in simulated scenarios of sex-imbalance in outcome prevalence and sex-imbalance in both representation and outcome prevalence.*** *Calibration plots of the models that predict 'abnormal' in scenarios of sex-imbalance in outcome prevalence and representation (simulation 3). A single calibration curve per condition and sex was generated by aggregating predicted probabilities and true outcomes across all five cross-validation folds. The smECE and calibration curves were then computed from these combined results. Panels show calibration plots grouped by outcome prevalence in the simulated scenarios;* ***A)*** *90% in women, 50% in men.* ***B)*** *50% in women, 10% in men.* ***C)*** *10% in women, 50% in men.* ***D)*** *10% in women, 90% in men. In panel A-D, Subplots in the top row show the calibration for men in the test sets, and in the bottom row the calibration for women. Subplots from left to right have the following proportion of women in training and validation datasets: 1.00, 0.75, 0.50, 0.25. 0.00.*

### Figure S2

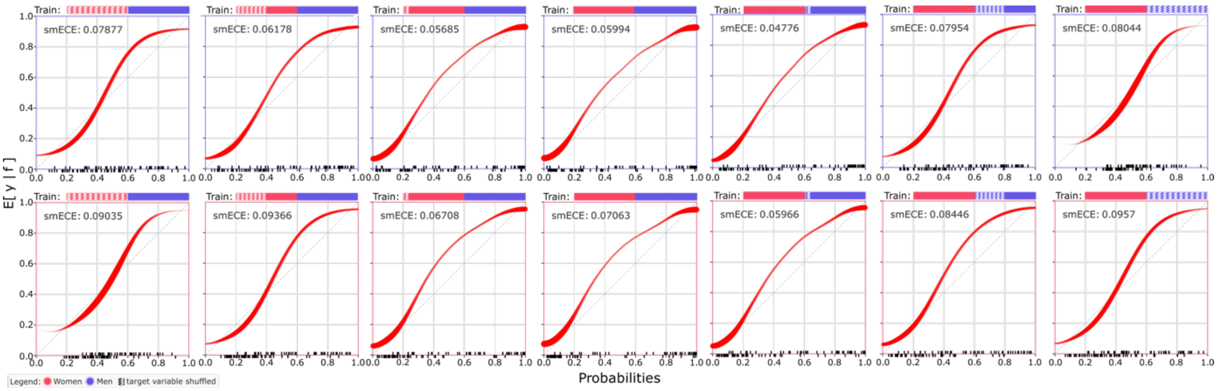


***Figure S2: Calibration in simulated scenarios of sex-dependent outcome misclassification.*** *Misclassifications were introduced by randomly shuffling certain percentages of the predicted outcomes in women or in men (simulation 4). A single calibration curve per condition and sex was generated by aggregating predicted probabilities and true outcomes across all five cross-validation folds. The smECE and calibration curves were then computed from these combined results. Subplots in the top row show the calibration for men in the test sets, and in the bottom row the calibration for women. Subplots from left to right have the following percentages of shuffled outcome (women:men): 100%:0%, 50%:0%, 10%: 0%, 0%:0% (reference), 0%:10%, 0%:50%, 0%:100%.*

### Figure S3

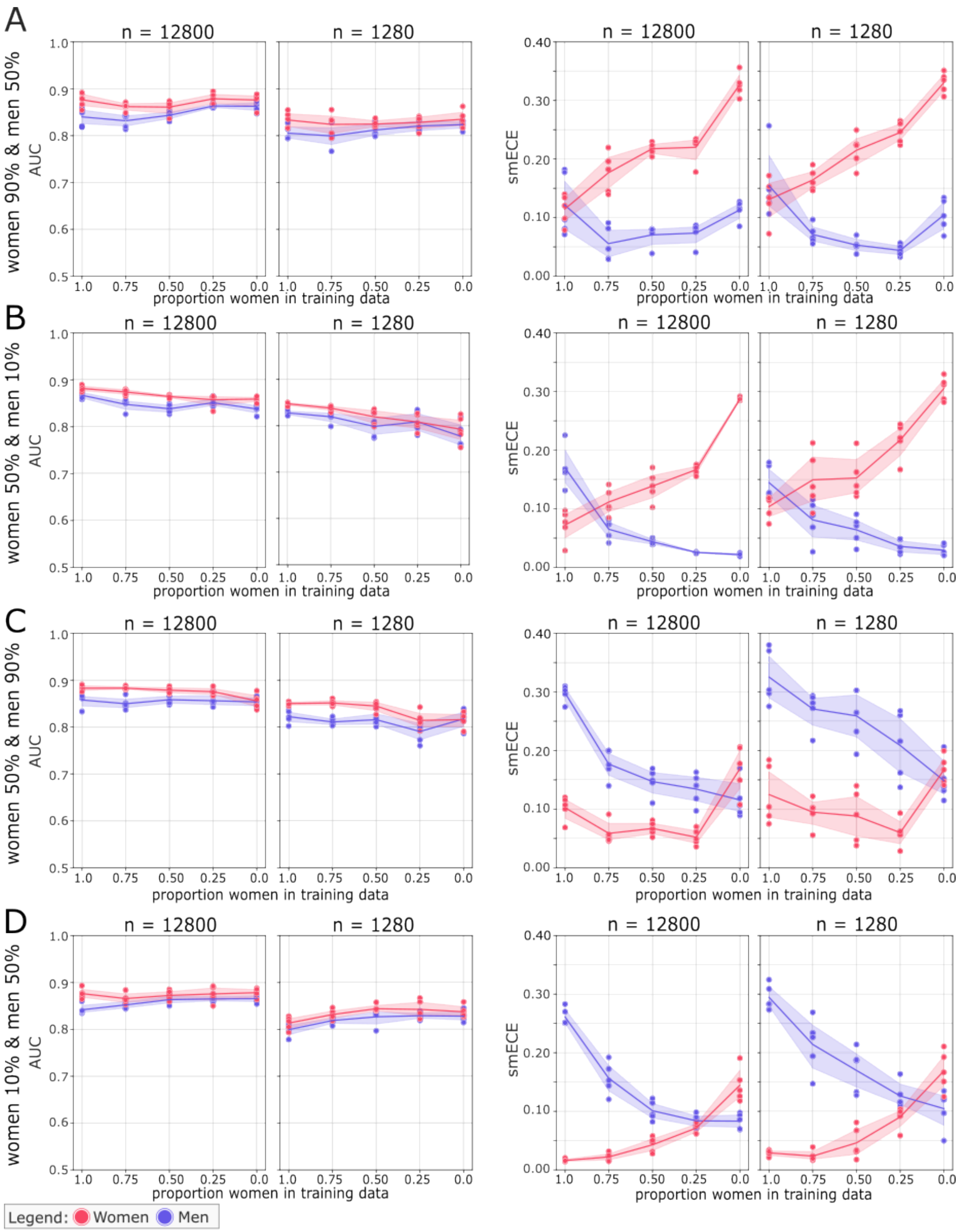


***Figure S3: Sex-stratified model performance examining the impact of sample size in simulated scenarios of sex-imbalance in both representation and outcome prevalence.***

*Using scenarios from simulation three, we depict model outcomes for two dataset sizes: a larger set ($n_{training} + n_{validation}$ = 12,800) and a smaller set ($n_{training} + n_{validation}$ = 1,280), with test set size held constant at around $n_{test}$ = 12,048. All models were trained to predict the outcome "abnormal". Each dot represents a single fold for either men or women; lines show mean performance and shaded areas indicate bootstrapped 95% confidence intervals. Panels show performance plots grouped by outcome prevalence in the simulated scenarios;* ***A)*** *90% in women, 50% in men.* ***B)*** *50% in women, 10% in men.* ***C)*** *10% in women, 50% in men.* ***D)*** *10% in women, 90% in men. In panel A-D, AUC and calibration error are shown for each scenario across both sample size conditions.*

### Figure S4

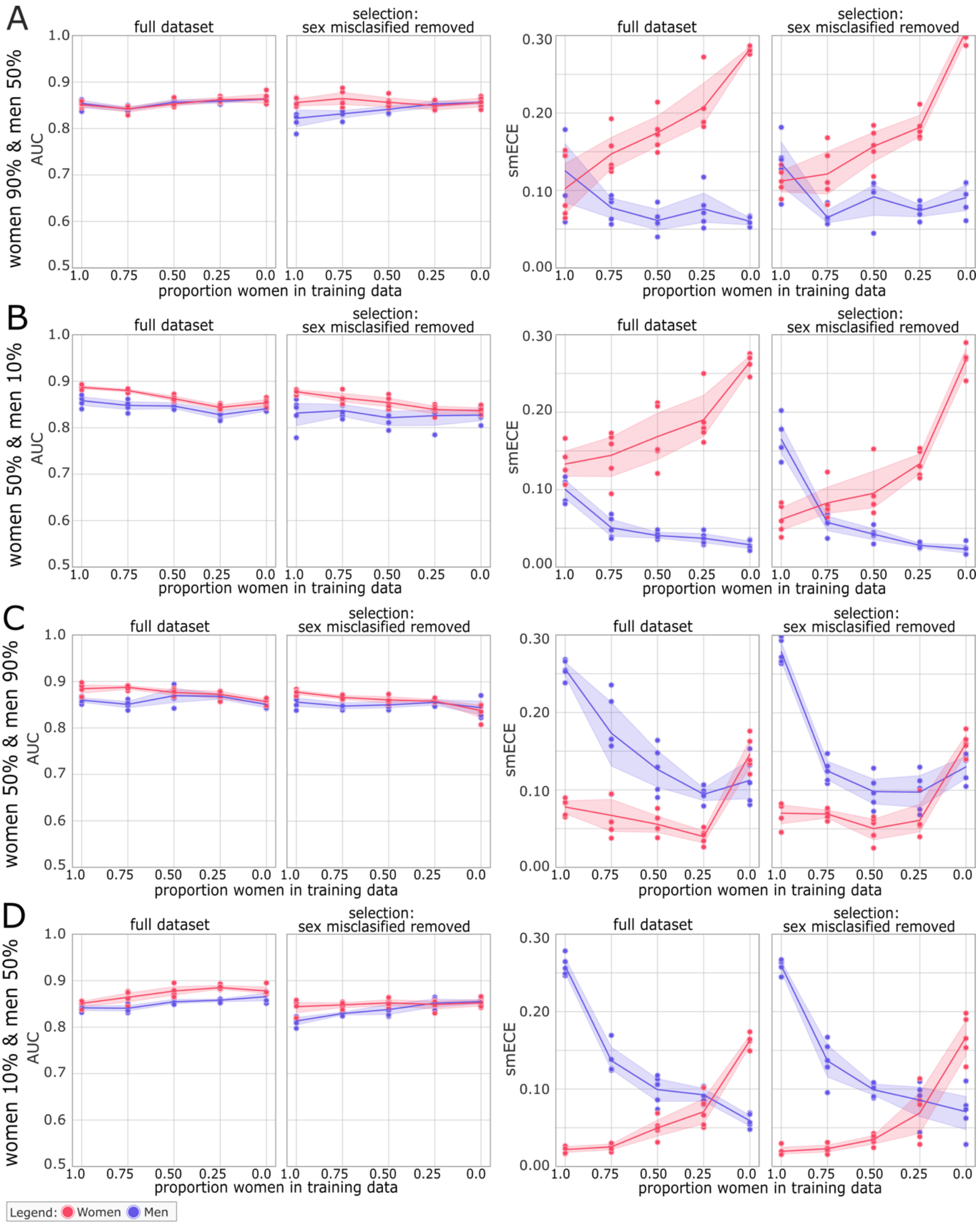

A
B
C
D
women 90% & men 50%
women 50% & men 10%
women 50% & men 90%
women 10% & men 50%
full dataset
selection:
sex misclasified removed
AUC
smECE
proportion women in training data
Legend: Women Men

***Figure S4: Sex-stratified model performance examining the impact of excluding patients with sex-misclassified ECGs from the training and validation data, in simulated scenarios of sex-imbalance in representation and outcome prevalence.*** *Shows model performance trained on the full datasets and those trained on subsets excluding patients whose sex was misclassified by a previously trained CNN model. The sample size of the test set remained consistent across all scenarios for the full dataset or the selected dataset. All models were trained to predict the outcome "abnormal". Each dot represents a single fold for either men or women; lines show mean performance and shaded areas indicate bootstrapped 95% confidence intervals. Panels show performance plots grouped by outcome prevalence in the simulated scenarios;* ***A)*** *90% in women, 50% in men.* ***B)*** *50% in women, 10% in men.* ***C)*** *10% in women, 50% in men.* ***D)*** *10% in women, 90% in men. In panel A-D, AUC and calibration error are shown for each scenario across both sample size conditions.*
